# Automated Variant Prioritization in Suspected Genetic Kidney Disease

**DOI:** 10.1101/2025.09.29.25336840

**Authors:** Nina Rank, Sören Lukassen, Manuel A. Anderegg, Kai-Uwe Eckardt, Jan P. Halbritter, Bernt Popp

## Abstract

**Introduction:** Despite the identification of >700 genes associated with inherited kidney disease, many affected individuals presumably remain genetically undiagnosed through testing of known disease genes. Novel disease gene discovery requires interpretation of large-scale sequencing data, which remains challenging.

**Methods:** We developed the Nephro Candidate Score (N-CS), a machine learning (ML) framework for variant prioritization combining a Nephro Gene Score (N-GS), Nephro Variant Score (N-VS), and heuristic Inheritance Score. The ML-based N-GS and N-VS predict gene-disease relevance and variant pathogenicity from diverse genomic features. Clinical utility was assessed in 1452 individuals with suspected inherited kidney disease using exome-wide candidate discovery and gene-level burden analysis, with external validation using recently proposed inherited kidney disease genes.

**Results:** The N-CS showed strong predictive performance, with the XGBoost-based N-GS achieving an AUC of 0.94 and the Logistic Regression-based N-VS an AUC of 0.99 on independent test sets. A gene set enrichment analysis demonstrated significant enrichment of kidney-related biological processes among top-ranked genes (p < 0.001). In our patient cohort, N-CS-guided variant filtering recovered established disease genes and highlighted *ERMP1* as a compelling candidate. Incorporating the N-GS in a polycystic kidney disease burden analysis reduced multiple-testing penalties, highlighting a moderate-effect candidate. Externally, the N-CS assigned high scores to 10 of 11 newly proposed kidney disease genes.

**Conclusion:** The N-CS is a digital framework to accelerate disease gene discovery and diagnosis. It is freely available as the NC-Scorer web tool (https://nc-scorer.kidney-genetics.org/) and a command-line interface enabling standardized, scalable candidate variant evaluation.

## Introduction

Chronic kidney disease (CKD) is a global health concern, affecting more than 10% of adults and is predicted to be the fifth leading cause of death worldwide by 2040^1^. Traditionally, hypertension and diabetes mellitus are considered responsible for two-thirds of CKD cases. However, this concept is largely based on clinical assumptions rather than histological confirmation, and hypertensive nephropathy has been questioned as a stand-alone disease entity^2^. With the integration of molecular genetics into CKD diagnostics, the contribution of monogenic and complex genetic causes has become increasingly recognized. At least 28% of patients with CKD have a positive family history^3,4^ - suggesting a genetic origin of the disease. Established monogenic etiologies, including Autosomal Dominant Polycystic Kidney Disease (ADPKD), Alport Syndrome and Congenital Anomalies of the Kidney and Urinary Tract (CAKUT) account for 30-50% of CKD in children and 10-30% in adults^5^. Despite the identification of >700 genes^6,7^ associated with rare and inherited kidney diseases, many patients remain without a molecular diagnosis, as conventional analyses fail to identify causal germline variants (e.g. 13% in ADPKD^8^, > 80% in CAKUT^9^). Identification of novel disease genes and pathogenic variants is an urgent medical need to avoid diagnostic odysseys, improve genetic counseling, and expand treatment options.

Advances in high-throughput sequencing, particularly exome sequencing, enable cost-effective genetic testing for individuals with unexplained CKD^10^. When diagnostic analysis of known genes is inconclusive, research-based variant filtering, evaluation, and interpretation become difficult due to the size of the necessary data sets. In principle, any rare variant that segregates with the clinical trait of interest may be disease-causing, but functional *in vitro* or *in vivo* investigation of all candidate variants is impossible. It is therefore necessary to prioritize and focus on the most promising variants. For this purpose, variants are often ranked manually by experts according to multiple criteria, including minor allele frequencies in large populations, inheritance mode, or the predicted effect on the encoded protein^11^. In addition, there is a variety of databases that provide information on the associated genes (e.g. on data from mouse models, protein-protein interactions, and RNA-seq data in general and specifically in the kidney). However, manually examining the various data sources for the entirety of candidate variants is extremely time-consuming, not scalable, and highly investigator-dependent. Therefore, there is a high need for an instrument that systematically and automatically prioritizes candidate variants for genetic kidney disease. Here, we present the Nephro Candidate Score (N-CS), a machine learning (ML) framework designed to address this challenge by providing a standardized, automated, and resource-efficient assessment of candidate variants.

## Methods

### Nephro Candidate Score

We developed the Nephro Candidate Score (N-CS) as a composite of three subscores, the Nephro Gene Score (N-GS), the Nephro Variant Score (N-VS), and the Inheritance Score (IS), incorporating properties of the corresponding gene, the variant itself, and its mode of inheritance. The N-CS ranges from 0.2 to 10 and the subscores are combined as follows:

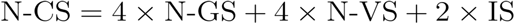

A schematic illustration of the N-CS components is depicted in Fig. 1.

**Fig. 1.**
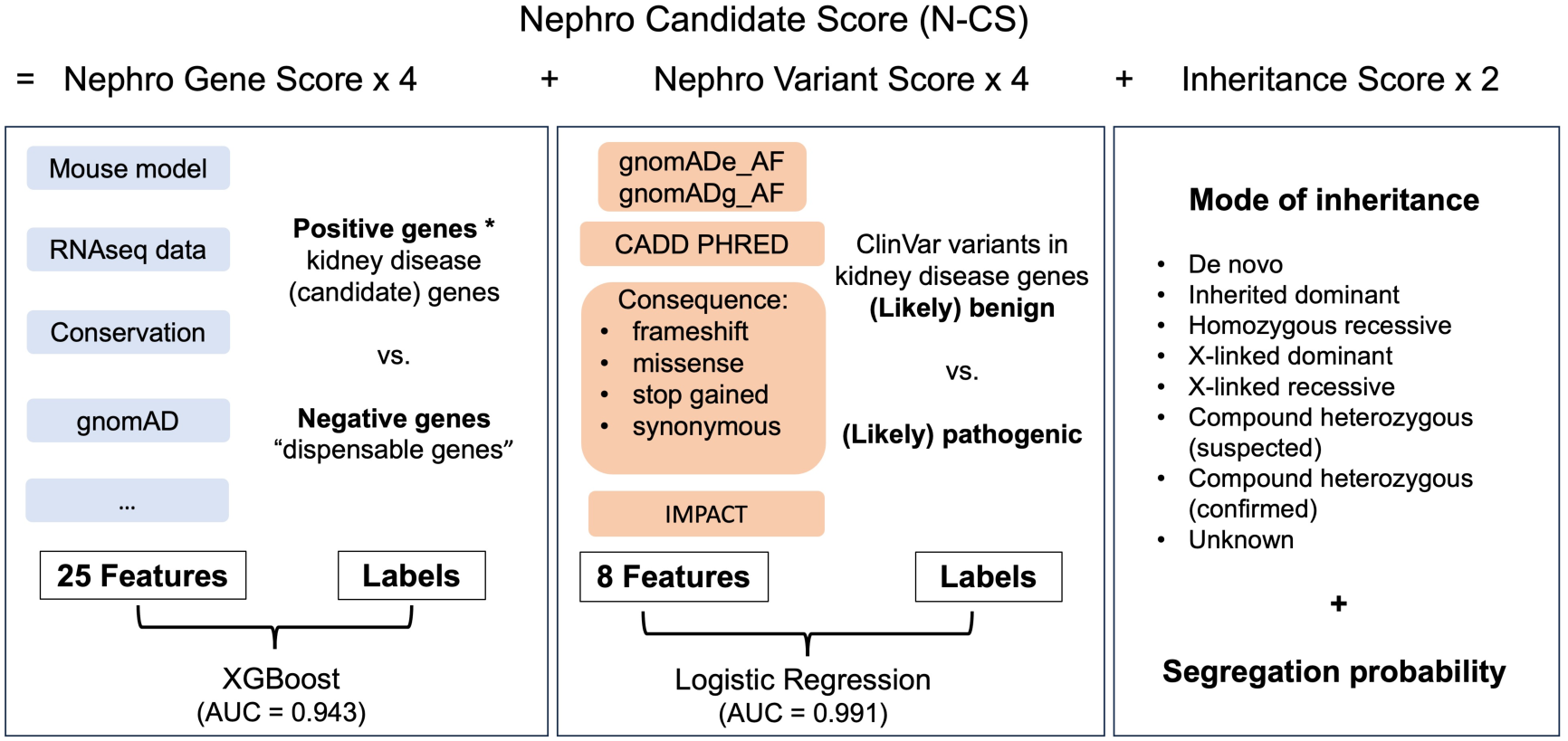
Components of the Nephro Candidate Score (N-CS).

### Nephro Gene Score

#### Model development

The N-GS is a ML-based model to classify genes as kidney disease-associated (positive) or not (negative). Positive Genes were defined as those with strong evidence for kidney disease involvement, sourced from the *Kidney-Genetics-v1* database^7^. We previously generated this publicly accessible resource that aggregates gene-disease associations from five streams of evidence: expert-curated lists, comprehensive literature reviews, clinical diagnostic panels, ontology-based searches in OMIM^12^, and automated literature extraction. Genes can reach a maximum evidence count (EC) of 5. Genes with EC ≥ 2 were considered positive, reflecting robust evidence supporting a gene-disease link. Negative Genes (“dispensable” genes) were defined as genes with homozygous loss-of-function variants in gnomAD^13,14^, no OMIM phenotype, and no Kidney-Genetics-v1 entry. The 109 features used in model training (Supplementary Methods, Table S1) include data from gnomAD, information on paralogues, promoter and exon CpG sites/conservation, mouse models, GTEx, and adult/fetal kidney single-cell DNA data. Twelve scikit-learn^15^ algorithms were trained (Supplementary Methods, Section S1). An XGBoost model using a reduced set of 25 features achieved the highest and most stable AUC and was selected as the final model. The N-GS ranges from 0 to 1, with higher scores indicating greater probability of kidney disease association.

#### Gene Set Enrichment Analysis

To further validate the model’s ability to identify kidney disease-associated genes, we performed a two-step Gene Set Enrichment Analysis (GSEA)^16,17^ (Supplementary Methods, Section S2) to assess whether genes involved in kidney-related biological processes were enriched among the top-ranked genes predicted by our model.

### Nephro Variant Score

The N-VS is a ML model that distinguishes pathogenic variants from benign variants. Training and test sets comprised all ClinVar^18^ variants in kidney disease-associated or candidate genes with an EC ≥ 2 in the Kidney-Genetics-v1 database. Only variants classified as “(Likely) Benign” and “(Likely) Pathogenic” were included. The initial feature set included 35 variant-level annotations derived using the Variant Effect Predictor (VEP)^19^ (Supplementary Methods, Table S2, Section S3). Decision trees and logistic regression were evaluated, both showing excellent performance. Logistic regression, using a reduced set of 8 features, was selected as the final model because it provides continuous probability estimates for integration into the N-CS. The N-VS ranges from 0 to 1, with higher values indicating a greater likelihood of pathogenicity.

### Inheritance Score

The IS quantifies a variant’s mode of inheritance by integrating inheritance pattern and family segregation data into a single metric. Each variant receives a base score reflecting its inheritance pattern, with higher scores assigned to patterns with stronger pathogenic potential (e.g. 0.95 for *de novo*, 0.1 for unknown inheritance). When segregation evidence is available, the score is boosted toward 1.0 according to the statistical strength of the segregation p-value. Conversely, when expected segregation evidence was missing, the score was reduced to reflect increased uncertainty. The final IS ranges from 0.1 to 1.0 (Supplementary Methods, Section S4).

### Web and Command-Line Implementations of the N-CS

To facilitate broad access and standardized variant assessment, we developed the Nephro Candidate Scorer (NC-Scorer), a user-friendly web tool available at https://nc-scorer.kidney-genetics.org/. The tool enables researchers and clinicians to apply the N-CS framework without requiring bioinformatics expertise and provides three primary search modes (Fig. 2):

● **Scoring Search:** Users can enter a single variant in Variant Call Format (VCF; e.g. `1-55051215-G-GA`) or transcript-based Human Genome Variation Society (HGVS) notation (e.g. `NM_001009944.3:c.11935C>T`). After selecting the inheritance mode, the tool calculates the composite N-CS and displays the individual components (N-GS, N-VS, and IS) with supporting annotations.
● **Variant Search:** Users can retrieve variant-specific information including the N-VS, the respective N-GS, and supporting evidence such as CADD scores and gnomAD population frequencies.
● **Gene Search:** Users can enter a gene symbol without further variant information to retrieve the N-GS.

**Fig. 2.**
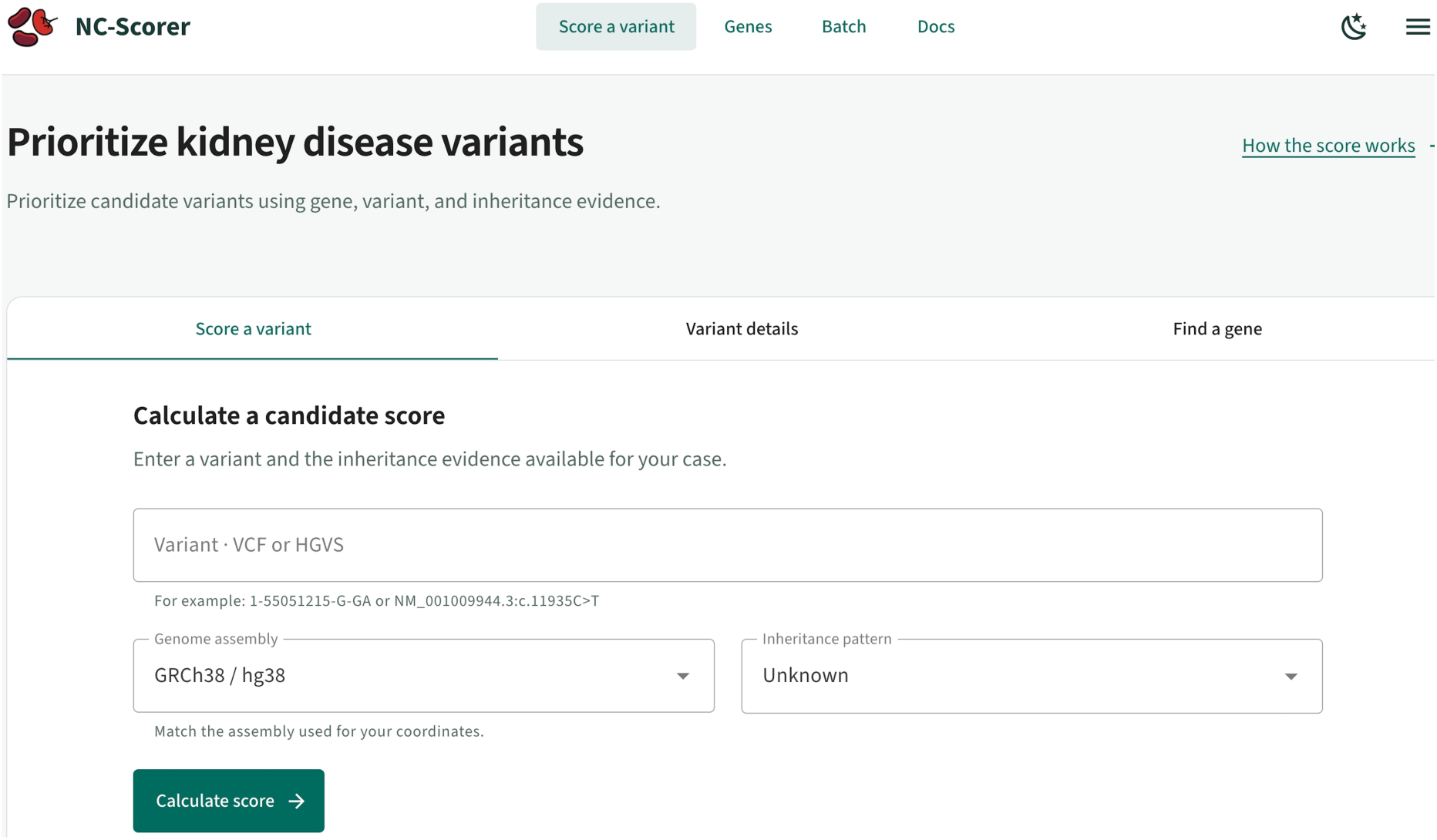
The NC-Scorer web interface. The tool provides a straightforward interface for calculating the Nephro Candidate Score (N-CS). Users can input a variant, specify the inheritance pattern, and receive a score. The platform also supports dedicated gene and variant searches, as well as batch processing.

Additional features include:

● **Batch scoring:** Enables simultaneous scoring of up to 200 variants, with results exportable in CSV/TSV, JSON, or VCF formats for downstream analyses and pipeline integration. Additional features include API caching to improve query speed.
● **Genes Score Overview**: Provides a sortable table of precomputed N-GS values for all genes, allowing users to browse genes ranked by kidney disease relevance.

For high-throughput applications, a command-line interface is also available. Both the web application and command-line interface are open-source and freely available.

### Application to real patient data

#### Study Cohort and Variant Identification

To evaluate the performance of the N-CS in real patient data, we applied the framework to a patient cohort of the Center of Rare Kidney Diseases (CeRKiD) at Charité – Universitätsmedizin Berlin, comprising 1452 individuals with suspected inherited kidney disease. All patients consented to the use of their data for scientific purposes. Clinical data and specific genetic data were obtained from patients enrolled in the CeRKiD study - approved by the Ethics Committee of Charite – Universitätsmedizin Berlin (approval number: EA4/066/21). All patients underwent whole-exome sequencing. Alignment to GRCh38 and single-sample variant calling with GATK HaplotypeCaller^20^ were performed on the Varvis platform (Limbus Medical Technologies). The resulting VCFs were uniformly hard-filtered and normalized with hardnormly (v0.7.3)^21^ and merged into a cohort-level multi-sample VCF. Variants were annotated with SnpEff and dbNSFP through sm-vcf-annotation (v1.2.0)^22^, then filtered and scored using VariantCentrifuge (v0.17.10)^23^ (Supplementary Methods, Section S5).

For the downstream analyses, we restricted our dataset to rare, high-confidence predicted loss-of-function (pLoF) variants by applying the following set of filters:

● Loss-of-function probability (pLoF) > 0.9
● Nonsense-mediated decay (NMD) escape probability > 0.9
● Predicted impact by SnpEff = HIGH
● Joint allele frequency (AF) < 0.01% in gnomAD v4.1
● Not classified as ‘benign’ in ClinVar

#### Experiment 1: NCS-Driven Discovery of High-Impact Rare Variant Burden

To evaluate the framework as an exome-wide discovery engine, we collapsed and counted qualifying rare variant gene-sample events across our cohort (see Supplementary Methods, Section S5.4 for details of event-clustering). Rather than using standard variant counts, analyses were restricted to genes with strong evidence of renal disease involvement or severe variant impact, defined as a N-CS ≥ 8. This hypothesis-free analysis had three objectives: (i) to confirm automated rediscovery of established, highly penetrant kidney disease genes (e.g., *PKD1, PKD2*); (ii) to identify potential phenotypic expansions of genes from the MorbidGenes panel^24^, not traditionally linked to CKD; and (iii) to nominate novel disease candidates.

#### Experiment 2: PKD Case-Control Association and NGS-Weighted False Discovery Rate Analysis

To assess enrichment of rare, high-impact variants in PKD, we performed an exome-wide, gene-level burden association analysis comparing patients with PKD and non-PKD controls using a two-tailed Fisher’s exact test. To increase statistical power, multiple testing correction was performed using a weighted False Discovery Rate (FDR) framework that incorporated prior clinical and biological knowledge through the N-GS. Raw gene weights *w_raw_* were derived from the N-GS as follows:

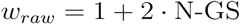

These weights were then globally normalized and used to reduce the multiple-testing burden for genes with prior evidence while preserving strict control of the overall false discovery rate (Supplementary Methods, Section S5.5).

### Validation on Novel Candidate Genes from Literature

To validate the N-CS’s ability to prioritize novel disease genes, we performed a systematic literature review using PubTator^25^ to identify candidate genes reported in 2024–2025 in high-impact genetics and nephrology journals (Supplementary Methods, Section S6). We identified 11 novel candidate genes that were not included in the N-CS model’s training set, comprising 46 reported variants. All variants were scored using the N-CS framework to evaluate its ability to independently prioritize newly reported gene–disease associations (Supplementary Results, Section S15).

## Results

### Nephro Gene Score

#### Performance and Distribution of the Nephro Gene Score

The final N-GS model is an XGBoost classifier using 25 features. It achieved an AUC of 0.943 on the independent test set. Additional performance metrics and feature importance are provided in the Supplementary Results, Sections S7-S8, Table S3, Fig. S1). Fig. 3a presents the distribution of the N-GS across dispensable genes, genes with an EC of 0-5, and "unassigned” genes (i.e., genes lacking both an EC and a classification as “dispensable”). As expected, dispensable genes received very low scores (median 0.03), whereas the median N-GS increased progressively with prior evidence (EC). Unassigned genes exhibited a broad, U-shaped distribution with a median score of 0.44 (Fig. 3b), suggesting two distinct subpopulations: one clustering near zero, likely representing genes unrelated to kidney disease, and another high-scoring group. Overall, 5,616 genes fall within the subpopulation with an N-GS ≥ 0.8. It is within this high-scoring subset that potential new candidate genes are most likely to be found, warranting further investigation, particularly if they harbor variants with high N-VS or IS.

**Fig. 3.**
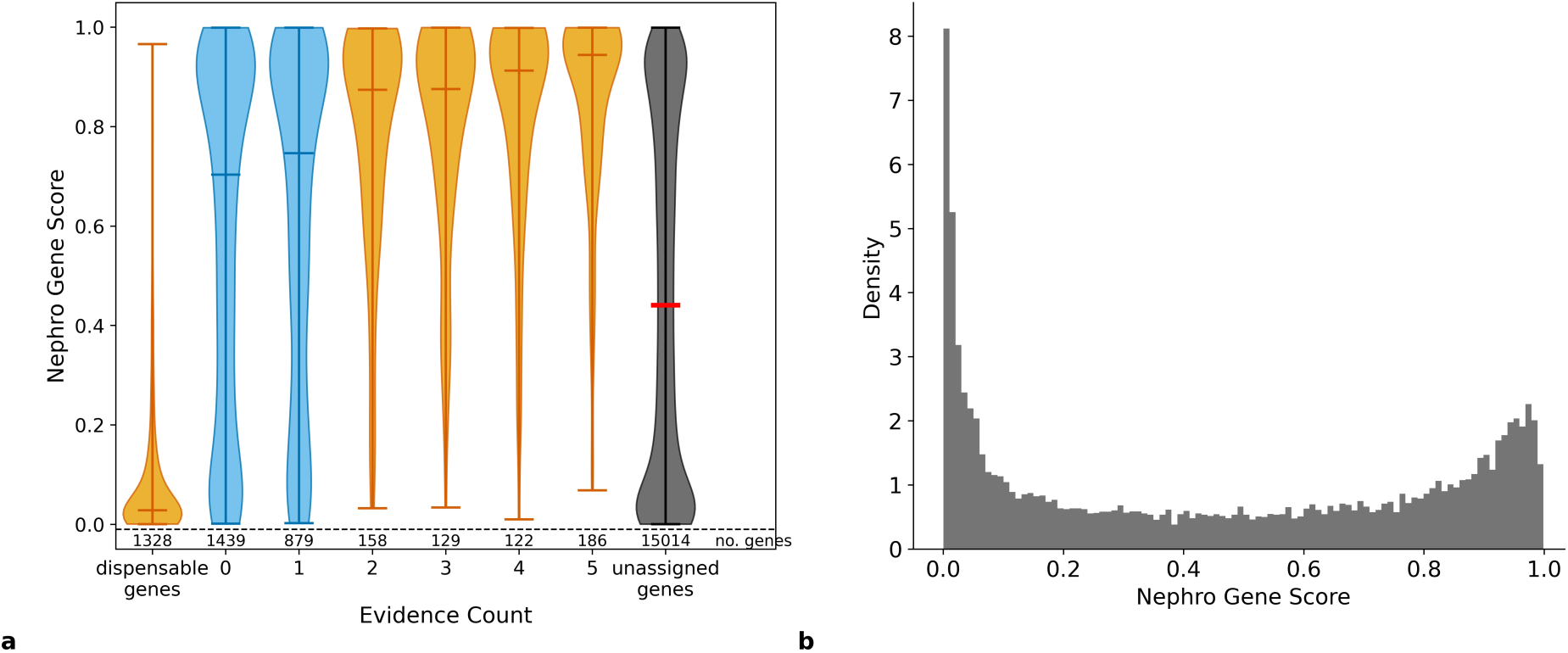
Distribution of Nephro Gene Scores (N-GS) across dispensable genes, genes with evidence counts (EC) 0–5, and unassigned genes (a) and distribution of the N-GS across unassigned genes (b). Unassigned genes do not have an evidence count, but are not classified as dispensable either. Yellow = genes used for ML training/testing, blue = genes with minor evidence for being kidney disease associated, grey = unassigned genes.

### Gene Set Enrichment Analysis

In the initial GSEA, 11 of the top 20 identified gene sets were linked to kidney-related GO terms. The top five ranked terms and the complete list of the top 20 ranked terms are provided in the Supplementary Results, Section S9, Fig. S2, Table S4. The subsequent enrichment analysis assessed whether the highest-ranked GO terms from the initial GSEA were specifically enriched for kidney-related biological processes. The kidney-GO-term set showed a highly significant enrichment, with a normalized enrichment score (NES) of 4.4 and a p-value of < 0.001.

### Nephro Variant Score

As the final N-VS model, we selected a logistic regression model with 8 features, achieving an AUC of 0.991 on the independent test set (for additional performance metrics, see Supplementary Results, Section S10, Table S5). SHAP analysis ranked the features by importance as follows: predicted functional impact, CADD-PHRED score, classification as a missense variant, gnomAD exome allele frequency (AF), classification as a synonymous variant, classification as a frameshift variant, gnomAD genome AF, and classification as a stop-gained variant (see Table S2 for feature descriptions). Including additional features yielded only a marginal AUC increase and was therefore omitted. Additionally, we evaluated the AUC across different functional impact groups (Table S6).

### Clinical Application in a Cohort with Suspected Inherited Kidney Diseases

#### Experiment 1: N-CS Driven Variant Filtering Recovers Known Kidney Disease Genes and Identifies Novel Candidate Genes

We assessed the exome-wide burden of rare predicted loss-of-function variants in 1452 individuals with suspected inherited kidney disease. To focus on variants with strong predicted disease relevance, we filtered for those with an N-CS ≥ 8. We then selected genes with at least three independent gene-sample events (Fig. 4). A gene-sample event was defined by collapsing adjacent variant rows within the same gene and sample into a single event if they were located within a 10-bp genomic window to account for complex alleles and multi-nucleotide variant clusters.

**Fig. 4.**
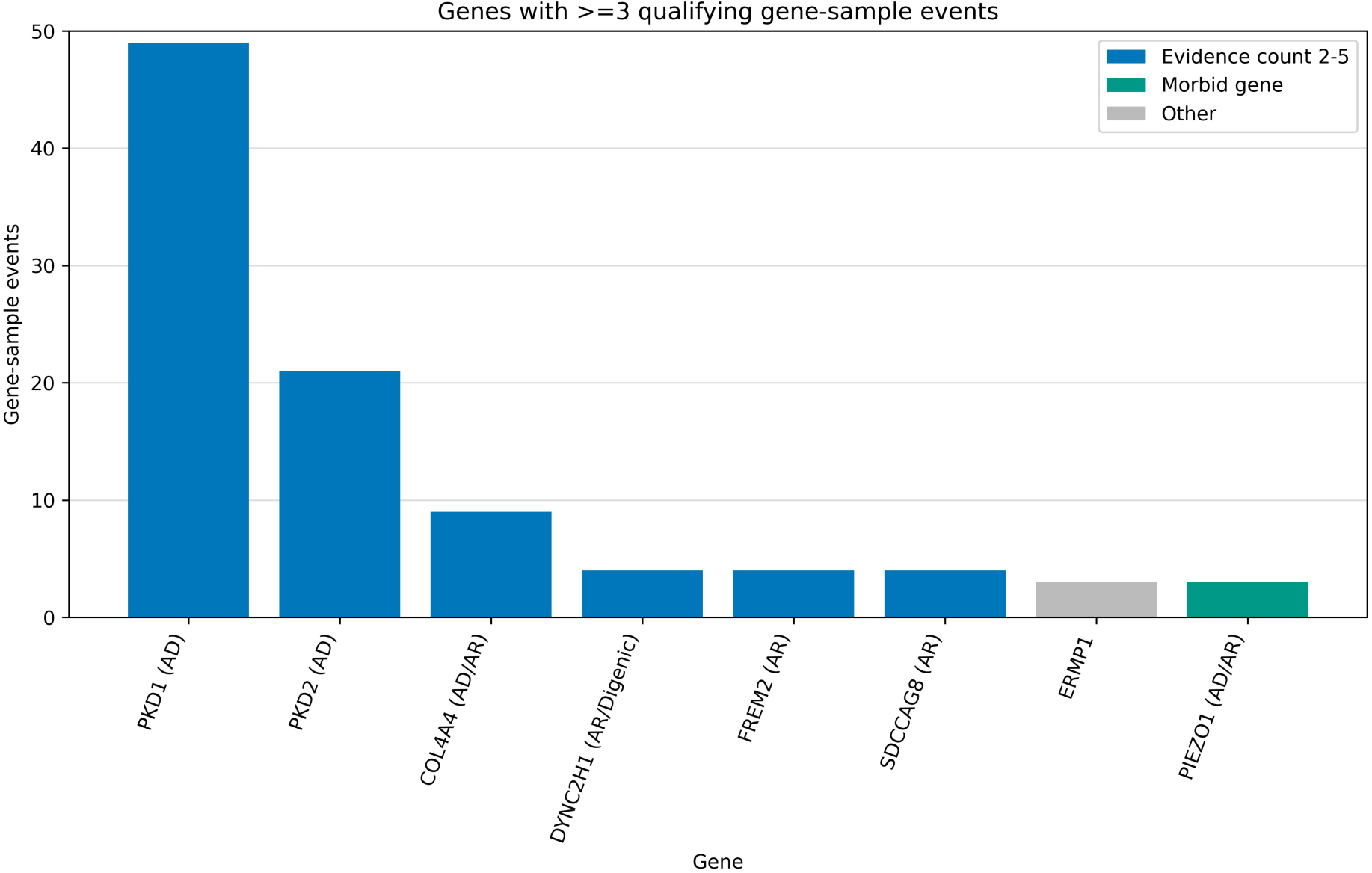
Exome-Wide Discovery of High-Impact Rare Variant Burden. Bar plot displaying candidate genes with a N-CS ≥ 8 that accumulated ≥ 3 qualifying independent gene-sample events across the study cohort. AD = autosomal dominant, AR = autosomal recessive. Blue = genes with an evidence count of ≥ 2 in the Kidney-Genetics-v1 database, green = genes from MorbidGenes panel, grey = uncharacterized genes.

The N-CS pipeline successfully rediscovered established kidney disease genes with *PKD1* (49 events) and *PKD2* (21 events) as the strongest signals, followed by *COL4A4* (9 events), causing Alport syndrome^26^, *SDCCAG8*^27^ (4 events), a recognized ciliopathy gene, *DYNC2H1* (4 events), which is associated with a syndromic ciliopathy with occasional cystic renal disease^28^, and the CAKUT-associated extracellular matrix gene *FREM2*^29^ (4 events).

Beyond these known disease genes, the pipeline prioritized *PIEZO1* (3 events), a MorbidGenes gene not previously linked to kidney disease, suggesting potential phenotypic expansion.

Additionally, the uncharacterized novel candidate *ERMP1* (Endoplasmic Reticulum Metallopeptidase 1) accumulated 3 gene-sample events. *ERMP1* currently lacks a characterized OMIM phenotype or an established link to genetic kidney disease, marking it as a prime, high-priority candidate for downstream functional validation studies. Clinical descriptions of affected individuals and the features driving the high N-CS are provided in the Supplementary Results, Section S11-12, Table S7, Fig. S3).

Omitting the N-CS filter markedly increased the number of candidate genes meeting the event threshold (Supplementary Results, Section S13, Fig. S4), demonstrating its value in reducing the candidate space while retaining the most biologically relevant disease genes.

#### Experiment 2: Integration of the N-GS Prior Refines Statistical Prioritization of Moderate-Effect Disease Drivers

To assess the enrichment of rare, high-impact variants in polycystic kidney disease (PKD), we performed a gene-level burden analysis comparing *n* = 405 PKD patients to *n* = 1047 non-PKD individuals with suspected inherited kidney disease (Fig. 5). After N-GS-weighted multiple-testing correction, the established PKD genes *PKD1* (p = 1.0 × 10^-41^, q_w_ = 4.9 × 10^-38^) and *PKD2* (p = 1.7 × 10^-19^, q_w_ = 3.9 × 10^-16^) were identified as the strongest exome-wide significant signals.

**Fig. 5.**
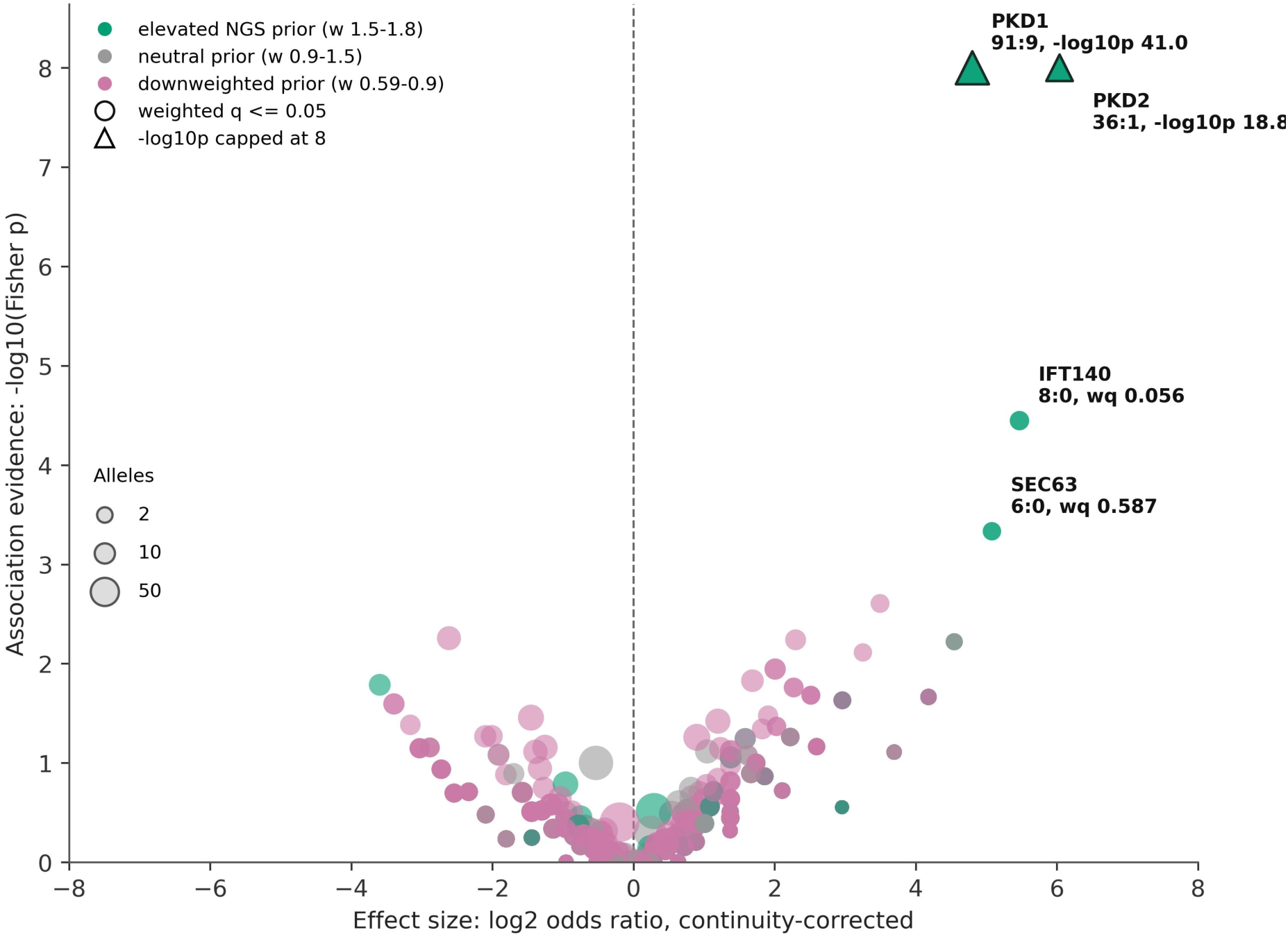
Exome-Wide Volcano Plot of PKD Burden Association Integrating an N-GS-Weighted Prior. Each data point represents a single evaluated gene. A rightward shift on the x-axis indicates enrichment in the PKD cohort. The effect of the N-GS prior applied during the weighted False Discovery Rate (FDR) correction is illustrated by color: green markers indicate an elevated prior (normalized weight 1.5–1.8) for genes with a high N-GS value, grey indicates a neutral baseline prior (normalized weight 0.9–1.5), and pink indicates a downweighted prior (normalized weight 0.59–0.9) penalizing genes with a low N-GS value. The size of each marker is proportional to the total number of qualifying variant alleles observed across the cohort.

Beyond these genes, incorporating the N-GS prior refined the statistical prioritization of secondary candidates by reducing the multiple-testing penalty according to biological knowledge. *IFT140* (8:0 case-to-control allele ratio, nominal p = 3.6 × 10^-5^, q = 0.098, q_w_ = 0.056) showed improved adjusted significance, approaching the formal exome-wide threshold with a weighted q-value of 0.056. *SEC63* ranked next (6:0 allele ratio, nominal p = 4.6 × 10^-4^, q = 0.959, q_w_ = 0.587); notably, these variants were detected in patients predominantly presenting with polycystic liver disease who had been broadly classified as PKD because no dedicated test panel for polycystic liver disease was available. Cohort metrics, continuity-corrected odds ratios, and rank-shift evaluations for all 52 nominally significant genes are detailed in Supplementary Results, Section S14, Fig. S5-6).

### The N-CS Prioritizes Novel Candidate Genes from Recent Literature

We evaluated the N-CS on a validation set of 46 variants from 11 recently reported kidney disease candidate genes (Supplementary Methods & Results, Sections S6,15, Table S8). These included candidates associated with CAKUT (*ARID3A*^30^*, NR6A1*^30,31^*, SOX13*^32^*, PHIP*^33^*, and TSHZ3*^34^*, PIEZO1*^35^), PKD (*UGGT1*^36^ *and CFAP47*^37^), ciliopathies (*WDR44*^38^), kidney dysplasia mimicking salt-wasting (*TFCP2L1*^39^), and proteinuric kidney disease (*MYO1C*^40^). The N-CS assigned scores ≥ 8 to the majority (9/11) candidate genes, with variants in 10 of the 11 novel genes receiving scores > 7, demonstrating effective prioritization of recently discovered disease genes across different inheritance modes (Fig. 6). Variants in *ARID3A*, *WDR44*, and *SOX13* consistently received high scores, whereas variants in *CFAP47* received comparatively lower scores. This was driven by a low gene-level prediction (N-GS = 0.03) — a consequence of a high gnomAD pLoF AF and a lack of kidney-specific expression— alongside weak variant-level evidence (N-VS), wherein two of the three missense mutations exhibited unremarkable CADD scores.

**Fig. 6.**
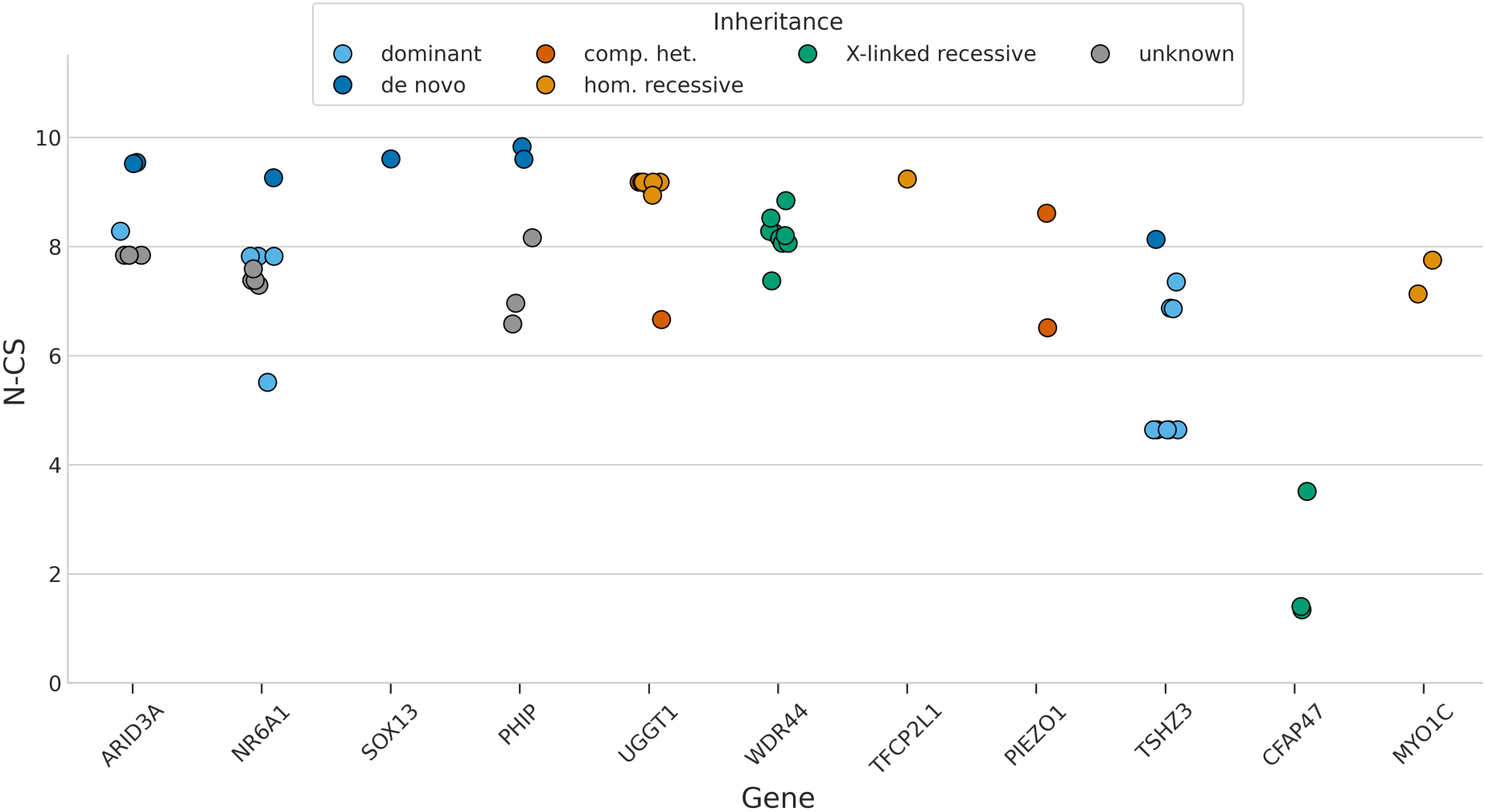
N-CS on novel candidate genes from a systematic literature review. The plot displays N-CS scores for 46 variants across 11 recently published candidate genes. Each point represents a single variant, colored by its mode of inheritance.

## Discussion

Current nephrogenetics faces the challenge of distinguishing causal variants from extensive background variation. This is particularly difficult in adults with suspected inherited kidney diseases, which are often characterized by late onset, incomplete penetrance, and variable expressivity. Together, these factors obscure clear genotype-phenotype correlations and hamper gene discovery. To address this challenge, we created and validated the N-CS, a ML-driven framework for automated candidate variant prioritization. By integrating gene-level biological relevance (N-GS), variant-level deleteriousness (N-VS), and inheritance mode (IS), the N-CS provides a standardized method for sifting through large-scale sequencing data.

While several computational tools support kidney genetics, the N-CS was developed to fill a specific niche. Inspired by the AutoCaSc framework previously developed for neurodevelopmental disorders,^41^ the N-CS was specifically designed for the evolutionary and clinical complexity of adult kidney diseases, where subtle pathogenic signals may be missed by rule-based approaches. By using supervised ML, the N-CS captures complex, non-linear relationships across 109 genomic features beyond human-defined heuristics.

Unlike tools such as *KidneyGPS*, which focuses on GWAS-based kidney function loci,^42^ the N-CS performs unbiased exome-wide prioritization without restriction to known genes. Compared with *KidneyNetwork*, which relies on kidney-specific co-expression patterns for gene discovery,^43^ the N-CS integrates diverse genomic evidence through a supervised model and generates a more granular, multi-dimensional score combining gene-level relevance (N- GS), variant-level deleteriousness (N-VS), and inheritance mode (IS). This provides a comprehensive framework for resolving individual genetic cases and identifying novel disease genes.

### Unbiased Exome-Wide Candidate Discovery

The exome-wide discovery analysis in individuals with suspected inherited kidney disease highlighted the ability of the N-CS framework to reduce a large search space to a focused set of candidates. After filtering for an N-CS ≥ 8, only 8 genes with ≥ 3 independent gene-sample events remained with PKD1, PKD2, and COL4A4 as the top-ranked signals, demonstrating that the framework can effectively recover established disease genes.

The capture of PIEZO1 highlights the potential of the N-CS to uncover hidden renal phenotypes and expand the spectrum of known disease genes. Although PIEZO1 has traditionally been associated with non-renal disorders, recent studies have linked biallelic loss-of-function variants to Prune Belly Syndrome with urinary tract abnormalities, supporting *PIEZO1* as a candidate gene for CAKUT^35^, and provided functional evidence for a role in kidney cystogenesis through calcium and cAMP signaling pathways^44^.

Highlighting our framework’s capacity for unbiased gene discovery, *ERMP1* emerged as a compelling novel candidate. Although it currently lacks an OMIM phenotype or established link to renal disease, recent findings support a potential role in kidney pathology. Correia de Sousa et al.^45^ found that endoplasmic reticulum metallopeptidase 1 (ERMP1) functions within the unfolded protein response to shield tubular epithelial cells from endoplasmic reticulum stress. Utilizing an *in silico* analysis of existing transcriptomic data, they showed that *ERMP1* is expressed across all major renal cell populations and exhibits upregulation in CKD. They also demonstrated that while complete *Ermp1* deletion in mice is lethal, heterozygous loss worsens age-associated chronic kidney damage. Additionally, they found that depleting *ERMP1* in human tubular cells drastically reduces metabolic function and cell survival. Together, these findings indicate that reduced *ERMP1* dosage impairs the kidney’s ability to withstand cellular stress, providing strong biological support for *ERMP1* as a candidate gene.

### Enhancing Statistical Power in Case-Control Associations

Exome-wide association analyses in adult-onset kidney disease are challenged by stringent multiple-testing penalties that can obscure moderate-effect signals. In our PKD association experiment, we incorporated the continuous N-GS biological prior into a weighted FDR framework to adjust this penalty according to gene-level evidence.

This approach improved the prioritization of biologically plausible candidates, including *IFT140,* a known ciliopathy gene, which approached exome-wide significance after weighting (q_w_ = 0.056 vs. unweighted q = 0.098). The lack of formal significance likely reflects limited cohort size (405 PKD cases). The foundational study by Senum et al. required screening approximately 1200 families—supplemented by massive national-scale datasets like the UK Biobank and the Genomics England 100000 Genomes Project—to confidently establish *IFT140* as a core ADPKD driver^46^. Seen in this light, the fact that the N-GS-weighted framework improved the adjusted significance of *IFT140* to the very cusp of the exome-wide threshold within a single, moderately sized clinical cohort underscores the framework’s statistical efficiency. By separating thousands of genes into low- and high-priority groups through ML-derived biological priors, the framework reduces the risk that genuine moderate-effect associations are lost to uniform multiple-testing correction.

### Clinical Utility and Accessibility

The N-CS demonstrated strong generalizability in our literature-based validation, effectively prioritizing variants from recently discovered CKD candidate genes and supporting its application beyond the original training data.

While primarily developed as a hypothesis-generation tool for research, the N-CS also has potential clinical utility by prioritizing rare variants of uncertain significance (VUS) identified through exome sequencing. By narrowing large numbers of candidate variants to a focused set for expert review and ACMG assessment, it may support more efficient diagnostic workflows and facilitate the resolution of challenging cases. Furthermore, its standardized scoring framework enables systematic re-analysis of existing sequencing cohorts using a consistent approach.

To promote broad adoption, we released the N-CS as an open-source toolkit, including a public web interface (https://nc-scorer.kidney-genetics.org/) and command-line implementation. This enables researchers with varying computational resources to apply, validate, and compare candidate prioritization approaches, supporting collaborative gene discovery and improved diagnostic efforts.

### Future Directions and Limitations

The current N-CS prioritizes coding variants, but could be extended to incorporate non-coding regulatory and structural variants by integrating additional functional genomic features, e.g. chromatin accessibility and regulatory interaction data. Disease-specific models for kidney disease subtypes, such as cystic kidney diseases, CAKUT, or glomerulopathies, may further improve specificity over the current pan-kidney score.

Integration with emerging AI approaches, such as LLMs, could enable automated literature synthesis and evaluation of biological plausibility for top-ranked candidates. Beyond causal variant discovery, the framework could help identify modifier genes influencing disease severity, progression, or extra-renal manifestations in monogenic kidney disorders, potentially explaining clinical variability.

This study has several limitations. Because the N-CS models were trained on currently known kidney disease genes and variants, they may be biased against genes involved in entirely novel biological mechanisms. Performance also depends on the quality and completeness of the underlying external databases. Furthermore, the current framework is limited to single-nucleotide variants and small indels and does not assess non-coding regulatory variants or larger structural variants.

## Conclusion

The N-CS provides an ML-based framework for prioritizing candidate variants in CKD by integrating gene-level evidence, variant-level pathogenicity, and inheritance patterns into a unified score. By addressing key challenges of adult-onset kidney disease, including late presentation and incomplete penetrance, it offers a standardized approach to accelerate gene discovery and support genetic diagnosis. As an openly available tool, the N-CS enables broad application by researchers and clinicians. Future validation in larger clinical cohorts will be essential to establish its clinical utility and generalizability.

## Disclosure

BP is employed by Labor Berlin - Charité Vivantes GmbH, a diagnostic laboratory offering clinical genetic testing for inherited kidney disease. Labor Berlin had no role in study design, data analysis, interpretation, or manuscript preparation. All other authors declare no competing interests.

## Software and Data Sharing

N-GS features were acquired and preprocessed in R (v4.3.2), while N-VS feature generation and all ML analyses were performed in Python (v3.7). The N-CS is released as open-source software: the NC-Scorer web application and command-line interface (https://github.com/halbritter-lab/nc-scorer), and the model training code and processed data (https://github.com/halbritter-lab/nephro_candidate_score). The underlying gene-disease evidence is at https://github.com/halbritter-lab/kidney-genetics-v1. Cohort processing used hardnormly (https://github.com/berntpopp/hardnormly), merge-multisample-vcf (https://github.com/scholl-lab/merge-multisample-vcf), sm-vcf-annotation (https://github.com/scholl-lab/sm-vcf-annotation) and VariantCentrifuge (https://github.com/scholl-lab/variantcentrifuge), all openly available. Exact versions, settings and annotation database releases for every component are given in Supplementary Methods, Section S5. Full individual exome sequencing data cannot be shared publicly due to patient privacy and data protection regulations.

## Data Availability

The N-CS is released as open-source software: the NC-Scorer web application and command-line interface (https://github.com/halbritter-lab/nc-scorer), and the model training code and processed data (https://github.com/halbritter-lab/nephro_candidate_score). The underlying gene-disease evidence is at https://github.com/halbritter-lab/kidney-genetics-v1. Cohort processing used hardnormly (https://github.com/berntpopp/hardnormly), merge-multisample-vcf (https://github.com/scholl-lab/merge-multisample-vcf), sm-vcf-annotation (https://github.com/scholl-lab/sm-vcf-annotation) and VariantCentrifuge (https://github.com/scholl-lab/variantcentrifuge), all openly available. Exact versions, settings and annotation database releases for every component are given in Supplementary Methods, Section S5. Full individual exome sequencing data cannot be shared publicly due to patient privacy and data protection regulations.

https://github.com/halbritter-lab/nc-scorer

https://github.com/halbritter-lab/kidney-genetics-v1

https://github.com/berntpopp/hardnormly

https://github.com/scholl-lab/merge-multisample-vcf

https://github.com/scholl-lab/sm-vcf-annotation

https://github.com/scholl-lab/variantcentrifuge

## Acknowledgements

JH receives funding from the German Research Foundation through the Heisenberg Programme (DFG, HA 6908/4-1). NR was funded by the Junior Digital Clinician Scientist Program of the Berlin Institute of Health (BIH) at Charité. MA was supported by the Swiss National Science Foundation through a Postdoc.Mobility fellowship (P500PM_222010).

## Authors‘ contributions

Conceptualization: NR, SL, MA, JH, BP Data curation: NR, BP

Formal Analysis: NR, BP Original draft: NR

Review & editing: NR, SL, MA, KE, JH, BP

## Declaration of generative AI and AI-assisted technologies in the manuscript preparation process

During the preparation of this work, the authors used ChatGPT and Gemini in order to assist with language editing, improving readability, and refining prose. After using these tools, the authors reviewed and edited the content as needed and take full responsibility for the content of the published article.

## Notes

### Author Declarations

To evaluate the performance of the N-CS in real patient data, we applied the framework to a patient cohort of the Center of Rare Kidney Diseases (CeRKiD) at Charitä - Universitätsmedizin Berlin, comprising 1452 individuals with suspected inherited kidney disease. All patients consented to the use of their data for scientific purposes. Clinical data and specific genetic data were obtained from patients enrolled in the CeRKiD study - approved by the Ethics Committee of Charitä - Universitätsmedizin Berlin (approval number: EA4/066/21).

### Summary of Updates

We have added a clinical application of the Nephro Candidate Score framework to a real world cohort of 1452 individuals with suspected inherited kidney disease. This new section includes an exome wide rare variant burden analysis for candidate discovery and a case control association study for polycystic kidney disease to further validate the clinical utility of our method.

## References

1. Foreman KJ, Marquez N, Dolgert A, et al. Forecasting life expectancy, years of life lost, and all-cause and cause-specific mortality for 250 causes of death: reference and alternative scenarios for 2016--40 for 195 countries and territories. Lancet. 2018;392(10159):2052–2090.

2. Freedman BI, Cohen AH. Hypertension-attributed nephropathy: what’s in a name? Nat Rev Nephrol. 2016;12(1):27–36.

3. Titze S, Schmid M, Köttgen A, et al. Disease burden and risk profile in referred patients with moderate chronic kidney disease: composition of the German Chronic Kidney Disease (GCKD) cohort. Nephrol Dial Transplant. 2015;30(3):441–451.

4. Zhang J, Thio CHL, Gansevoort RT, Snieder H. Familial aggregation of CKD and heritability of kidney biomarkers in the general population: The Lifelines Cohort Study. Am J Kidney Dis. 2021;77(6):869–878.

5. KDIGO Conference Participants. Genetics in chronic kidney disease: conclusions from a Kidney Disease: Improving Global Outcomes (KDIGO) Controversies Conference. Kidney Int. 2022;101(6):1126–1141.

6. Groopman E, Goldstein D, Gharavi A. Diagnostic utility of exome sequencing for kidney disease. Reply. N Engl J Med. 2019;380(21):2080–2081.

7. Popp, B. Kidney-Genetics-v1: Kidney-Genetics - Database of Kidney-Related Genes. Github. Accessed September 7, 2025. https://github.com/halbritter-lab/kidney-genetics-v1

8. Schönauer R, Baatz S, Nemitz-Kliemchen M, et al. Matching clinical and genetic diagnoses in autosomal dominant polycystic kidney disease reveals novel phenocopies and potential candidate genes. Genet Med. 2020;22(8):1374–1383.

9. van der Ven AT, Vivante A, Hildebrandt F. Novel Insights into the Pathogenesis of Monogenic Congenital Anomalies of the Kidney and Urinary Tract. J Am Soc Nephrol. 2018;29(1):36–50.

10. Halbritter J, Figueres L, Van Eerde AM, et al. Chronic Kidney Disease of unexplained cause (CKDx): a consensus statement by the Genes & Kidney Working Group of the ERA. Nephrol Dial Transplant. Published online June 3, 2025. doi:10.1093/ndt/gfaf092

11. Richards S, Aziz N, Bale S, et al. Standards and guidelines for the interpretation of sequence variants: a joint consensus recommendation of the American College of Medical Genetics and Genomics and the Association for Molecular Pathology. Genet Med. 2015;17(5):405–424.

12. Amberger JS, Bocchini CA, Schiettecatte F, Scott AF, Hamosh A. OMIM.org: Online Mendelian Inheritance in Man (OMIM®), an online catalog of human genes and genetic disorders. Nucleic Acids Res. 2015;43(Database issue):D789–D798.

13. Karczewski KJ, Francioli LC, Tiao G, et al. The mutational constraint spectrum quantified from variation in 141,456 humans. Nature. 2020;581(7809):434–443.

14. Leitão E, Schröder C, Parenti I, et al. Systematic analysis and prediction of genes associated with monogenic disorders on human chromosome X. Nat Commun. 2022;13(1):6570.

15. Pedregosa F, Varoquaux G, Gramfort A, et al. Scikit-learn: Machine Learning in Python. J Mach Learn Res. 2011;12(Oct):2825–2830.

16. Subramanian A, Tamayo P, Mootha VK, et al. Gene set enrichment analysis: a knowledge-based approach for interpreting genome-wide expression profiles. Proc Natl Acad Sci U S A. 2005;102(43):15545–15550.

17. Fang Z, Liu X, Peltz G. GSEApy: a comprehensive package for performing gene set enrichment analysis in Python. Bioinformatics. 2023;39(1). doi:10.1093/bioinformatics/btac757

18. Landrum MJ, Lee JM, Riley GR, et al. ClinVar: public archive of relationships among sequence variation and human phenotype. Nucleic Acids Res. 2014;42(Database issue):D980-D985.

19. McLaren W, Gil L, Hunt SE, et al. The Ensembl Variant Effect Predictor. Genome Biol. 2016;17(1):122.

20. McKenna A, Hanna M, Banks E, et al. The Genome Analysis Toolkit: a MapReduce framework for analyzing next-generation DNA sequencing data. Genome Res. 2010;20(9):1297–1303.

21. Popp B. Hardnormly. Github. Accessed May 29, 2026. https://github.com/berntpopp/hardnormly

22. Popp B. Sm-Vcf-Annotation. Github. Accessed May 29, 2026. https://github.com/scholl-lab/sm-vcf-annotation

23. Popp B. Variantcentrifuge: A Pipeline for Filtering Annotated Variant Call Format Files. Github. Accessed May 29, 2026. https://github.com/scholl-lab/variantcentrifuge

24. Jauss RT, Popp B, Bachmann J, Jamra RA, Platzer K. The MorbidGenes panel: a monthly updated list of diagnostically relevant rare disease genes derived from diverse sources. medRxiv. Published online April 15, 2024. doi:10.1101/2024.04.15.24305833

25. Wei CH, Allot A, Lai PT, et al. PubTator 3.0: an AI-powered literature resource for unlocking biomedical knowledge. Nucleic Acids Res. 2024;52(W1):W540–W546.

26. Mochizuki T, Lemmink HH, Mariyama M, et al. Identification of mutations in the alpha 3(IV) and alpha 4(IV) collagen genes in autosomal recessive Alport syndrome. Nat Genet. 1994;8(1):77–81.

27. Otto EA, Hurd TW, Airik R, et al. Candidate exome capture identifies mutation of SDCCAG8 as the cause of a retinal-renal ciliopathy. Nat Genet. 2010;42(10):840–850.

28. Dagoneau N, Goulet M, Geneviève D, et al. DYNC2H1 mutations cause asphyxiating thoracic dystrophy and short rib-polydactyly syndrome, type III. Am J Hum Genet. 2009;84(5):706–711.

29. Kohl S, Hwang DY, Dworschak GC, et al. Mild recessive mutations in six Fraser syndrome-related genes cause isolated congenital anomalies of the kidney and urinary tract. J Am Soc Nephrol. 2014;25(9):1917–1922.

30. Milo Rasouly H, Krishna Murthy SB, Vena N, et al. Exome analysis links kidney malformations to developmental disorders and reveals causal genes. Nat Commun. 2025;16(1):7290.

31. Neelathi UM, Ullah E, George A, et al. Variants in NR6A1 cause a novel oculo vertebral renal syndrome. Nat Commun. 2025;16(1):6111.

32. Merz LM, Kolvenbach CM, Wang C, et al. Trio exome sequencing identifies de novo variants in novel candidate genes in 19.62% of CAKUT families. Genet Med. 2025;27(7):101432.

33. Rivera-Munoz EA, Zhao XE, Rosenfeld JA, et al. Clinical exome sequencing efficacy and phenotypic expansions involving non-isolated congenital anomalies of kidney and urinary tract (CAKUT+). Eur J Hum Genet. Published online September 6, 2025. doi:10.1038/s41431-025-01929-3

34. Kesdiren E, Martens H, Brand F, et al. Heterozygous variants in the teashirt zinc finger homeobox 3 (TSHZ3) gene in human congenital anomalies of the kidney and urinary tract. Eur J Hum Genet. 2025;33(1):44–55.

35. Amado NG, Nosyreva ED, Thompson D, et al. PIEZO1 loss-of-function compound heterozygous mutations in the rare congenital human disorder Prune Belly Syndrome. Nat Commun. 2024;15(1):339.

36. Dardas Z, Harrold L, Calame DG, et al. Bi-allelic UGGT1 variants cause a congenital disorder of glycosylation. Am J Hum Genet. 2025;112(5):1139–1157.

37. Mori T, Fujimaru T, Liu C, et al. CFAP47 is implicated in X-linked polycystic kidney disease. Kidney Int Rep. 2024;9(12):3580–3591.

38. Accogli A, Shakya S, Yang T, et al. Variants in the WDR44 WD40-repeat domain cause a spectrum of ciliopathy by impairing ciliogenesis initiation. Nat Commun. 2024;15(1):365.

39. Vaqueiro Graña M, Madariaga L, Gómez-Conde S, Iceta Lizarraga A, Hualde Olascoaga J, Ariceta G. Ultra-rare severe kidney dysplasia mimicking salt-wasting tubulopathy associated with TFCP2L1 gene variants. Pediatr Nephrol. Published online June 26, 2025. doi:10.1007/s00467-025-06804-3

40. Elmubarak I, Shril S, Mansour B, et al. Recessive variants in MYO1C as a potential novel cause of proteinuric kidney disease. Pediatr Nephrol. 2024;39(10):2939–2945.

41. Lieberwirth JK, Büttner B, Klöckner C, Platzer K, Popp B, Abou Jamra R. AutoCaSc: Prioritizing candidate genes for neurodevelopmental disorders. Hum Mutat. 2022;43(12):1795–1807.

42. Stanzick KJ, Stark KJ, Gorski M, et al. KidneyGPS: a user-friendly web application to help prioritize kidney function genes and variants based on evidence from genome-wide association studies. BMC Bioinformatics. 2023;24(1):355.

43. Boulogne F, Claus L, Wiersma H, et al. KidneyNetwork: Using kidney-derived gene expression data to predict and prioritize novel genes involved in kidney disease. doi:10.21203/rs.3.rs-1870632/v1

44. Fan Q, Hadla M, Peterson Z, et al. Activation of PIEZO1 attenuates kidney cystogenesis in vitro and ex vivo. Kidney360. 2024;5(11):1601–1612.

45. Correia de Sousa M, Arnoux G, Yvon R, et al. ERMP1 as a newly identified endoplasmic reticulum stress gatekeeper in chronic kidney disease. Am J Physiol Renal Physiol. 2025;328(3):F375–F388.

46. Senum SR, Li YSM, Benson KA, et al. Monoallelic IFT140 pathogenic variants are an important cause of the autosomal dominant polycystic kidney-spectrum phenotype. Am J Hum Genet. 2022;109(1):136–156.

