## Supplementary material for "Automated Variant Prioritization in Suspected Genetic Kidney Disease"

#### Supplementary Methods

##### Section S1: Model development of the N-GS

For the N-GS, we compiled 109 features from open-source platforms (Table S1). The annotated gene set was split into a 80% training and 20% test set. We trained 12 scikit-learn[1] machine learning algorithms to classify genes into kidney-associated and not kidney-associated genes (XGBClassifier, AdaBoostClassifier, DecisionTreeClassifier, RandomForestClassifier, SVC, LinearSVC, GaussianNB, MLPClassifier, QuadraticDiscriminantAnalysis, GaussianProcessClassifier, RidgeClassifier, KNeighborsClassifier). The hyperparameters were tuned using GridSearch and 5-fold cross-validation on the training set, prioritizing configurations that achieved both a high and stable AUC across slight variations.

**Table S1. Candidate feature groups considered for training of the N-GS model.**

| Feature group/Feature names | Description | Source | No. of feat. | Selected for the final model |
|---|---|---|---|---|
| ssRNA-seq data of the adult kidney | Single cell RNA-seq data from the adult kidney | [2] [3] | 26 | |
| CL_0000115_pc, CL_0000653_pc, CL_0002306_pc, CL_1000692_pc, CL_1000768_pc, CL_1000849_pc, CL_1001106_pc, CL_1001107_pc, CL_1001111_pc, CL_1001318_pc, CL_1001431_pc, CL_1001432_pc, CL_1000452_pc | Percentage of cells expressing the gene in the following cell types: endothelial cell, podocyte, epithelial cell of the proximal tubule, kidney interstitial fibroblast, kidney connecting tubule epithelial cell kidney distal convoluted tubule epithelial cell, kidney loop of Henle thick ascending limb epithelial cell, kidney loop of Henle thin ascending limb epithelial cell, kidney loop of Henle thin descending limb epithelial cell, renal interstitial pericyte, kidney collecting duct principal cell, kidney collecting duct intercalated cell parietal epithelial cell | | 13 | no |
| CL_0000115_me, CL_0000653_me, CL_0002306_me, CL_1000692_me, CL_1000768_me, CL_1000849_me, CL_1001106_me, CL_1001107_me, CL_1001111_me, CL_1001318_me, CL_1001431_me, | Gene expression value in the following cell types: endothelial cell, podocyte, epithelial cell of the proximal tubule, kidney interstitial fibroblast, kidney connecting tubule epithelial cell kidney distal convoluted tubule epithelial cell, kidney loop of Henle thick ascending limb epithelial cell, | | 13 | CL_1001432_me - gene expression value in kidney collecting duct intercalated cell |

| | | | | |
|---|---|---|---|---|
| CL_1001432_me,<br>CL_1000452_me | kidney loop of Henle thin ascending limb epithelial cell,<br>kidney loop of Henle thin descending limb epithelial cell,<br>renal interstitial pericyte,<br>kidney collecting duct principal cell,<br>kidney collecting duct intercalated cell<br>parietal epithelial cell | | | |
| ssRNA-seq data of the fetal kidney | Single cell RNA-seq data from the fetal kidney | [4] | 11 | |
| mesangial_perc_expr,<br>metanephric_perc_expr,<br>ureteric_bud_perc_expr,<br>stromal_perc_expr,<br>vascular_endothelial_perc_expr | Percentage of mesangial, metanephric, ureteric bud, stromal, and vascular endothelial kidney cells expressing the respective gene | | 5 | no |
| mesangial_nTPM,<br>metanephric_nTPM,<br>ureteric_bud_nTPM,<br>stromal_nTPM,<br>vascular_endothelial_nTPM | Quantile-normalized TPM for mesangial, metanephric, ureteric bud, stromal, and vascular endothelial kidney cells | | 5 | no |
| fetal_kidney_tau | Tissue specificity index (τ) for the fetal kidney, calculated using the method of Yanai et al. [5] | | 1 | yes |
| gnomAD data | gnomAD-derived constraint metrics | [6] | 34 | |
| gnomad_obs_mis | Observed number of missense variants in gnomAD. | | 1 | no |
| gnomad_exp_mis | Expected number of missense variants in gnomAD. | | 1 | no |
| gnomad_oe_mis | Observed-to-expected ratio of missense variants in gnomAD. | | 1 | yes |
| gnomad_mu_mis | Mutation rate for missense variants in gnomAD. | | 1 | no |
| gnomad_possible_mis | Possible number of missense variants based on sequence context in gnomAD. | | 1 | no |
| gnomad_obs_mis_pphen | Observed number of missense variants predicted to be damaging (polyphen) in gnomAD. | | 1 | no |
| gnomad_exp_mis_pphen | Expected number of missense variants predicted to be damaging (polyphen) in gnomAD. | | 1 | no |
| gnomad_oe_mis_pphen | Observed-to-expected ratio of damaging missense variants (polyphen) in gnomAD. | | 1 | yes |
| gnomad_possible_mis_pphen | Possible number of damaging missense variants (polyphen) in gnomAD. | | 1 | yes |
| gnomad_obs_syn | Observed number of synonymous variants in gnomAD. | | 1 | no |
| gnomad_exp_syn | Expected number of synonymous variants in gnomAD. | | 1 | no |
| gnomad_oe_syn | Observed-to-expected ratio of synonymous variants in gnomAD. | | 1 | yes |

| | | | | |
|---|---|---|---|---|
| gnomad_mu_syn | Mutation rate for synonymous variants in gnomAD. | | 1 | no |
| gnomad_possible_syn | Possible number of synonymous variants in gnomAD. | | 1 | no |
| gnomad_obs_lof | Observed number of loss-of-function (LoF) variants in gnomAD. | | 1 | no |
| gnomad_mu_lof | Mutation rate for loss-of-function variants in gnomAD. | | 1 | no |
| gnomad_possible_lof | Possible number of loss-of-function variants in gnomAD. | | 1 | no |
| gnomad_exp_lof | Expected number of loss-of-function variants in gnomAD. | | 1 | no |
| gnomad_pLI | Probability of a gene being loss-of-function intolerant (pLI) in gnomAD. | | 1 | no |
| gnomad_pRec | Probability of a gene being recessive loss-of-function tolerant in gnomAD. | | 1 | no |
| gnomad_pNull | Probability of a gene being loss-of-function tolerant in gnomAD. | | 1 | no |
| gnomad_oe_lof | Observed-to-expected ratio of loss-of-function variants in gnomAD. | | 1 | no |
| gnomad_syn_z | Z-score for the deviation of synonymous variants from the expected number in gnomAD. | | 1 | yes |
| gnomad_mis_z | Z-score for the deviation of missense variants from the expected number in gnomAD. | | 1 | yes |
| gnomad_lof_z | Z-score for the deviation of loss-of-function variants from the expected number in gnomAD. | | 1 | no |
| gnomad_oe_lof_upper_rank | Upper bound rank of the observed-to-expected ratio for loss-of-function variants in gnomAD. | | 1 | no |
| gnomad_n_sites | Total number of sites analyzed in the gene in gnomAD. | | 1 | yes |
| gnomad_classic_caf | Cumulative allele frequency of variants in gnomAD. | | 1 | yes |
| gnomad_max_af | Maximum allele frequency of variants in gnomAD. | | 1 | yes |
| gnomad_p | Probability score associated with observed variant distributions in gnomAD. | | 1 | no |
| gnomad_exp_hom_lof | Expected number of homozygous loss-of-function variants in gnomAD. | | 1 | no |
| gnomad_cds_length | Coding sequence length of the gene in gnomAD. | | 1 | no |
| gnomad_num_coding_exons | Number of coding exons in the gene based on gnomAD. | | 1 | no |

| | | | | |
|---|---|---|---|---|
| gnomad_gene_length | Total length of the gene, including non-coding regions, in gnomAD. | | 1 | no |
| GTEx data | Bulk RNA expression data across multiple human tissues | [7] | 30 | |
| gtex_adipose_tissue, gtex_adrenal_gland, gtex_breast, gtex_cervix, gtex_colon, gtex_endometrium, gtex_esophagus, gtex_fallopian_tube, gtex_heart_muscle, gtex_kidney, gtex_liver, gtex_lung, gtex_ovary, gtex_pancreas, gtex_pituitary_gland, gtex_prostate, gtex_retina, gtex_salivary_gland, gtex_skeletal_muscle, gtex_skin, gtex_small_intestine, gtex_spinal_cord, gtex_spleen, gtex_stomach, gtex_testis, gtex_thyroid_gland, gtex_urinary_bladder, gtex_vaginal, gtex_brain_median | Normalized TPM across multiple human tissues (adrenal gland, breast, cervix, colon, endometrium, esophagus, fallopian tube, heart muscle, kidney, liver, lung, ovary, pancreas, pituitary gland, prostate, retina, salivary gland, skeletal muscle, skin, small intestine, spinal cord, spleen, stomach, testis, thyroid gland, urinary bladder, vagina), Median normalized TPM across brain tissues | | 29 | gtex_adipose_tissue, gtex_adrenal_gland, gtex_kidney, gtex_liver, gtex_retina, gtex_salivary_gland, gtex_pancreas, gtex_testis, gtex_esophagus |
| gtex_tau | Normalized tissue specificity index (τ) calculated using the method of Yanai et al. [5] | | 1 | no |
| Ensembl Biomart v 109 | Number of paralogues, observed-to-expected CpG ratios and conservation scores | [8] | 7 | |
| no_paralogues_95, no_paralogues_85, no_paralogues_75 | Number of close paralogues above the 95th, 85th, and 75th percentiles (Target %ID and Query %ID) | | 3 | no_paralogues_75 |
| prom_CpG_o2e_ratio, avg_exon_CpG_o2e_ratio | Observed-to-expected CpG ratio of the promoter region and exons of the respective gene | | 2 | prom_CpG_o2e_ratio |
| avg_phasCons_exons, avg_phasCons_promoter | Average conservation PhastCons scores of coding exons and the promoter region of the respective gene | | 2 | yes |
| Mouse Genome Informatics (MGI) data | Mouse model data | [9] | 1 | |
| max_mgi_kid | Association of mouse genotypes (homozygous or heterozygous knock-out) with the renal/urinary system phenotype (MPO term "MP:0005367") or its children, annotated with human orthologs | | 1 | yes |

#### Section S2: Gene Set Enrichment Analysis of the N-GS

We employed a two-step Gene Set Enrichment Analysis (GSEA)[10,11] to assess whether our model's top-ranked genes were functionally linked to kidney-related biological processes. First, we constructed a set of Gene Ontology (GO) terms relevant to renal processes, referred to as “kidney-GO-terms.” This set included the terms GO:0003014 (“renal system process”) and GO:0072001 (“renal system development”) and all their descendants, along with their associated genes, derived from the "GO_Biological_Process_2023" dataset in the Enrichr database[12]. Next, we performed an initial GSEA using the full “GO_Biological_Process_2023” gene set collection. In this analysis, genes were ranked according to their predicted N-GS. The GSEA algorithm then assessed whether specific gene sets were enriched among genes with higher N-GS rankings, providing a Normalized Enrichment Score (NES) for each gene set. A second enrichment analysis tested whether kidney-GO-terms were preferentially enriched among the top-ranked GO terms based on the NES values from the first GSEA. This step yielded a single NES value for the kidney-GO-terms set, referred to as the "observed NES". To assess the statistical significance of this enrichment, we repeated the procedure 1,000 times, each time randomly shuffling the N-GS values among the genes in the first GSEA. This generated a null distribution of 1,000 NES values, against which we compared the observed NES to determine its significance.

#### Section S3: Feature Preprocessing, Dataset Stratification, and Sampling Strategy for the N-VS Model Training

For development of the N-VS, the 35 variant annotations shown in Table S2 were derived using the Variant Effect Predictor (VEP) tool[13].

During exploratory analysis, the *IMPACT* feature - a classification of a variant's predicted functional severity on a gene or protein developed by Ensembl[14] - was identified as a highly significant determinant of variant classification. To enhance the model’s ability to distinguish between the *LOW* and *MODERATE* IMPACT groups - representing the two middle classes in the hierarchy (MODIFIER, LOW, MODERATE, HIGH) - the training was performed on a dataset with balanced IMPACT groups. For this purpose, the complete variant dataset was first split into an 80% subset and a 20% subset. From the 80% subset, a training set with balanced IMPACT groups was created. An independent test set, maintaining the original distribution of IMPACT groups, was then sampled from the remaining data, which included the 20% subset as well as the variants from the 80% subset that were not used in the training set. Missing values in the dataset were imputed using the median values from the respective features in the training set. Prior to model training, all features were standardized using standard scaling to improve convergence speed and ensure numerical stability. For feature importance analysis, we employed SHAP (SHapley Additive exPlanations)[15]. As the final model, we chose a logistic regression model with a reduced set of 8 features. Including more features only marginally increased the AUC, with the reduced model remaining more interpretable and efficient without sacrificing performance.

**Table S2. Candidate features considered for training of the N-VS model.** In cases where multiple values were associated with a variant due to the presence of multiple transcripts, the method outlined in the third column was applied.

| Feature | Description | Value Selection for Multiple Transcript Values | Ref | Selected for the final model |
|---|---|---|---|---|
| CONSEQUENCE | Type of genetic alteration (e.g., missense variant, frameshift variant, synonymous variant). | Binary features were created for all possible consequences, with a value of 1 assigned if any transcript associated with the variant included the specific consequence. | [16] | synonymous variant<br>missense variant<br>frameshift variant<br>stop gained |
| IMPACT | Predicted severity of the effect of the variant. Values: MODIFIER, LOW, MODERATE, HIGH | The most severe impact was selected with priority given in the order: HIGH > MODERATE > LOW > MODIFIER | [14] | yes |
| GNOMADE_AF | Allele frequency of the variant in the gnomAD exome dataset. | The single available value, if not NA, was used. | [6] | yes |
| GNOMADG_AF | Allele frequency of the variant in the gnomAD genome dataset. | The single available value, if not NA, was used. | [6] | yes |
| CADD_PHRED | CADD score that predicts the deleteriousness of the variant on a scale from 0 to 99. | The single available value, if not NA, was used. | [17] | yes |
| NMD | Indicates whether the variant is subject to nonsense-mediated decay (NMD). | 1 if any transcript had a value of 1. | [14] | no |

| Table S2. Candidate features considered for training of the N-VS model. In cases where multiple values were associated with a variant due to the presence of multiple transcripts, the method outlined in the third column was applied. | | | | |
|---|---|---|---|---|
| **Feature** | **Description** | **Value Selection for Multiple Transcript Values** | **Ref** | **Selected for the final model** |
| SPLICEAI_PRED_DS_AG, SPLICEAI_PRED_DS_AL, SPLICEAI_PRED_DS_DG, SPLICEAI_PRED_DS_DL | Delta scores for acceptor (AG, AL) and donor (DG, DL) splice sites predicted by SpliceAI; the single available value was used if not NA | The single available value, if not NA, was used. | [18] | no |
| PHASTCONS100WAY_VERTEBRATE_RANKSCORE | Conservation score across 100 vertebrate species, indicating evolutionary conservation of the variant. | The single available value, if not NA, was used. | [19] | no |

#### Section S4: Derivation of the Inheritance Score

To quantify the strength of evidence for a variant's mode of inheritance, we developed the Inheritance Score (IS). This score systematically evaluates the observed inheritance pattern and adjusts for the presence or absence of statistical support from segregation analysis. Each variant was first assigned a base score based on its presumed inheritance pattern, reflecting the a priori significance of each mode of transmission. The scores were assigned as follows: 0.95 for *de novo*; 0.8 for homozygous, compound heterozygous, or autosomal recessive; 0.7 for X-linked recessive; 0.5 for X-linked dominant; 0.4 for autosomal dominant or possible compound heterozygous; and 0.1 for patterns classified as unknown. If a segregation p-value was available, the base score was adjusted upward toward 1.0 according to the strength of evidence. Specifically, a negative-log transformation of the p-value was scaled relative to a threshold value ($\gamma = 0.001$), such that p-values $\leq 0.001$ provided maximal support. This ensured that stronger segregation evidence boosted the score closer to 1.0, while absent or neutral evidence ($p = 1$) left the score unchanged from the base. If no segregation p-value was provided and the inheritance pattern was one for which segregation evidence would normally be expected, such as autosomal dominant, autosomal recessive, X-linked, homozygous, or compound heterozygous, a penalty factor was applied to reflect this missing evidence. Specifically, the adjusted score was reduced by 20% under these conditions. Patterns exempt from this requirement, such as *de novo* or unknown, were not penalized when segregation data were absent. The final score was therefore calculated as

$$IS = \begin{cases} \left[ baseScore + (1 - baseScore) \times \min\left(1, \frac{-\log(\max(p,\epsilon))}{-\log(\gamma)}\right) \right] \times 0.8, & \text{if segregation expected but missing} \\ baseScore + (1 - baseScore) \times \min\left(1, \frac{-\log(\max(p,\epsilon))}{-\log(\gamma)}\right), & \text{otherwise} \end{cases}$$

The Inheritance Score thus ranges from 0.1 for variants with an unknown pattern up to 0.95 by default for high-confidence *de novo* variants, but can approach 1.0 in the presence of strong segregation evidence.

#### Section S5: Bioinformatics Pipeline and Variant Filtering of the CeRKiD study data

##### Section S5.1. VCF Normalization and Alignment Processing

Exome sequencing and primary data analysis for the CeRKiD cohort were performed at Labor Berlin - Charite Vivantes GmbH in the course of routine diagnostic testing. Reads were aligned to GRCh38 and variants were called per sample with GATK HaplotypeCaller[20] on the Varvis platform (Limbus Medical Technologies GmbH, Rostock, Germany), which returned one filtered, VEP-annotated VCF per sample. Analyses were retrieved programmatically from the diagnostic instance using varvis-download (v0.29.0)[21]. No re-alignment or re-calling from raw reads was performed for this study, and all downstream analyses start from these 1452 single-sample VCFs. These VCFs were subsequently standardized using the automated variant normalization framework hardnormly (v0.7.3)[22] against the GRCh38 no-alt analysis set (GCA_000001405.15). Multi-allelic sites were split and variants uniformly left-aligned, GATK-recommended hard-filter thresholds were applied to the non-allele-specific quality metrics FS, MQ, SOR, ReadPosRankSum and MQRankSum, and genotype-level filters flagged heterozygous calls with a read depth below 10 or a variant allele fraction outside 0.2 to 0.8 and homozygous calls with a read depth below 5. The FILTER labels supplied with the diagnostic VCFs were retained and the study-wide filters appended alongside them, so that downstream analysis could be restricted to records passing both (Section S5.3). Processing was restricted to the cohort's specific exome target capture regions, taken as the union of the three capture kits represented in the cohort (287,314 merged regions) with 100 bp of padding. Crucially, to maximize sensitivity in genomic loci known to harbor highly complex sequence duplications (such as *PKD1*), conventional hard-exclusion intervals were explicitly bypassed during initial filtering. Instead, these complex genomic regions were preserved and carried downstream strictly as review annotations rather than hard filters. The 1452 normalized single-sample VCFs were then combined into one cohort-level multi-sample VCF using merge-multisample-vcf (v0.2.0)[23], a Snakemake workflow wrapping bcftools[24]. Merging proceeded in two stages, eight batches of up to 200 samples followed by a single final merge, with allele counts, frequencies and genotype missingness recomputed after each stage. This multi-sample VCF is the input to the annotation described in Section S5.2.

##### Section S5.2. Cohort-Wide Annotation and Software Environment

Annotation and filtering were separate stages. Filtering used VariantCentrifuge (v0.17.10)[25] and is described in Section S5.3. The merged multi-sample VCF was annotated by the Snakemake-based[26] workflow sm-vcf-annotation (v1.2.0[27]), which scatters annotation across 100 genomic intervals spanning the canonical contigs and concatenates the results. Functional consequences were assigned with SnpEff against the GRCh38.p14 database with loss-of-function annotation enabled and a splice-region intron window of 12 bp. Transcript- and variant-level annotations were integrated from the following genomic resources:

- **Transcript Framework**: MANE (Matched Annotation from NCBI and EMBL-EBI)[28] Select RefSeq transcript definitions.
- **Functional & Splice Metrics**: SnpEff[29] annotations capturing predicted functional impact (IMPACT), loss-of-function probability (LOF), and nonsense-mediated decay escape (NMD).
- **Pathogenicity & Population Frequency**: The dbNSFP v5.2a[30,31] profile, integrating global joint allele frequencies (AF) and allele counts (AC) from gnomAD v4.1[6], alongside updated ClinVar[32] clinical significance registries (ClinVar_CLNSIG), HGMD Pro 2021.4[33] variant class (hgmd_CLASS), and splice predictions from dbscSNV v1.1[34] and SPIDEX v1.0[35]. dbscSNV and SPIDEX are distributed on hg19 and were converted to GRCh38 by liftover, so their coordinates are inherited rather than native and variants that failed conversion carry no splice score. All annotations reported in this study were generated by this pipeline; the VEP annotations supplied with the diagnostic VCFs were removed during normalization and were not used.
- **Technical Quality Context**: Genome in a Bottle (GIAB)/NIST[36] GRCh38 v3.0 technical-region BED files mapping low-complexity zones, including GIAB_TANDEM_REPEAT, GIAB_SIMPLE_REPEAT, GIAB_HOMOPOLYMER_LOW_COMPLEXITY, GIAB_SEGMENTAL_DUPLICATION, and GIAB_LOW_MAPPABILITY.

##### Section S5.3. Downstream Multi-Stage VariantCentrifuge Filtering

High-throughput pre-filtering of the global VCF was completed using bcftools to restrict downstream analysis to premium-quality calls executing standard pass conditions (FILTER="PASS" AND QUAL > 300 AND INFO/AC[0] < 15). SnpEff consequence splitting was executed prior to applying variant selection criteria.

To enrich for high-impact alleles, VariantCentrifuge isolated rare, non-benign variants matching an expanded biological loss-of-function heuristic defined as:

1. Loss-of-function probability (pLoF) > 0.9
2. Nonsense-mediated decay escape probability > 0.9
3. Predicted impact by SnpEff = HIGH
4. Joint allele frequency (AF) < 0.01% in gnomAD v4.1
5. Not classified as ‘benign’ in ClinVar consensus labels within either ClinVar or dbNSFP tracking modules.

**Section S5.4. Quality Control, Post-Processing, and Event Collapsing**

For our first experiment, the N-CS-driven discovery of high-impact rare variant burden, specialized clustering filters were applied during exploratory analysis to counter local multi-nucleotide variant (MNV) inflation and technical misrepresentations within highly homologous regions:

- **Adjacent Row Collapse**: Co-occurring adjacent variant rows identified within the same individual sample, residing on the same chromosome, and positioned within 10 base pairs (bp) of one another were programmatically collapsed into a single gene-sample event.
- **Low-Complexity Repeat Adjustments**: Variants localized within low-complexity tracks (GIAB_TANDEM_REPEAT, GIAB_SIMPLE_REPEAT, or GIAB_HOMOPOLYMER_LOW_COMPLEXITY) were explicitly excluded from final gene-sample event-count summaries.
- **Complex Loci Preservation**: Segmental duplications or low mappability regions (e.g., *PKD1* exons 1–33) were explicitly *exempt* from hard drop-filters during visualization to prevent the loss of genuine pathogenic variations in complex kidney disease drivers.
- **Technical Artifact Blacklisting**: To ensure the integrity of candidate discovery, a manual review was performed on top-scoring genes. A figure-level artifact blacklist was applied to systematically exclude genes whose high scoring signal was strictly dominated by technical artifacts or representation errors arising from poor mapping quality.

##### Section S5.5: Mathematical Framework for NGS-Weighted FDR and Association Adjustments

To integrate domain-specific knowledge into the exome-wide association analysis without inflating the false positive rate, raw weights calculated via the N-GS formula ( $w_{raw} = 1 + 2 \cdot \text{N-GS}$) were normalized across all m tested genes to ensure the mean weight equals 1:

$$\frac{1}{m}\sum_{i=1}^{m} w_i = 1$$

The nominal Fisher's exact p-values ($P_i$) for each gene were then adjusted into weighted p-values ($P^*_i$) by dividing by their normalized weight:

$$P_i^* = \frac{P_i}{w_i}$$

The standard Benjamini-Hochberg (BH) procedure was subsequently applied to these ordered, weighted p-values to generate the final adjusted q-values.

Additionally, during the calculation of effect sizes ($log_2$-odds ratios), a Haldane-Anscombe continuity correction was applied to handle mathematical anomalies (such as infinite values) in candidate genes with zero variant counts in either the case or control groups. A constant value of +0.5 was programmatically added to all cells of the 2x2 contingency table before calculating the final $log_2$-odds ratio for visualization.

#### Section S6: Literature Search Strategy for Validation Set

To create an independent validation set of novel candidate genes, a systematic literature search was performed using the PubTator[37] interface. The objective was to identify genes associated with inherited kidney disease that were published recently and thus not included in the N-CS training data.

The search was conducted using the following comprehensive query:

```
(kidney OR renal OR CAKUT OR urinary OR nephro OR CKD OR ESRD) AND
(gene OR genetic OR candidate OR syndrome)
```

The results were filtered to include publications from the years 2024 and 2025. To focus on high-impact findings, the search was restricted to the following leading journals in genetics and nephrology: *Nature Genetics*, *Nature Communications*, *Genetics in Medicine*, *European Journal of Human Genetics*, *American Journal of Human Genetics*, *Journal of the American Society of Nephrology*, *Kidney International*, *Pediatric Nephrology*, and *Kidney International Reports*. To ensure our validation set consisted of truly novel or emerging candidates, we excluded any gene with an evidence count > 1 in the *Kidney-Genetics* database. This final filtering step resulted in the identification of the 11 novel candidate genes and 46 associated variants.

### Supplementary Results

#### Section S7: Model Performance of the N-GS

Table S3 presents the performance metrics of the N-GS in both the training and the test set:

**Table S3. Performance metrics of the Nephro Gene Score across the training and test set**.The threshold dependent metrics were yielded based on a threshold of 0.5 for positive/negative classification.

| | Training set | Test set |
|---|---|---|
| **Area-under-curve (AUC)** | 0.994 | 0.943 |
| **Precsion-Recall-AUC** | 0.988 | 0.892 |
| **Accuracy** | 0.968 | 0.877 |
| **Sensitivity** | 0.924 | 0.798 |
| **Specificity** | 0.988 | 0.911 |
| **False-discovery-rate** | 0.029 | 0.202 |
| **False-positive-rate** | 0.012 | 0.089 |
| **Negative predictive value** | 0.967 | 0.911 |
| **Positive predictive value** | 0.971 | 0.798 |

#### Section S8: Feature importance in the N-GS model

We used SHAP to analyze the feature importance of the N-GS model. A beeswarm plot is depicted in Fig. S1. It illustrates the feature's importance on the model's predictions. Each dot represents a gene with higher absolute values indicating a greater influence of a feature on the N-GS prediction for the respective gene.

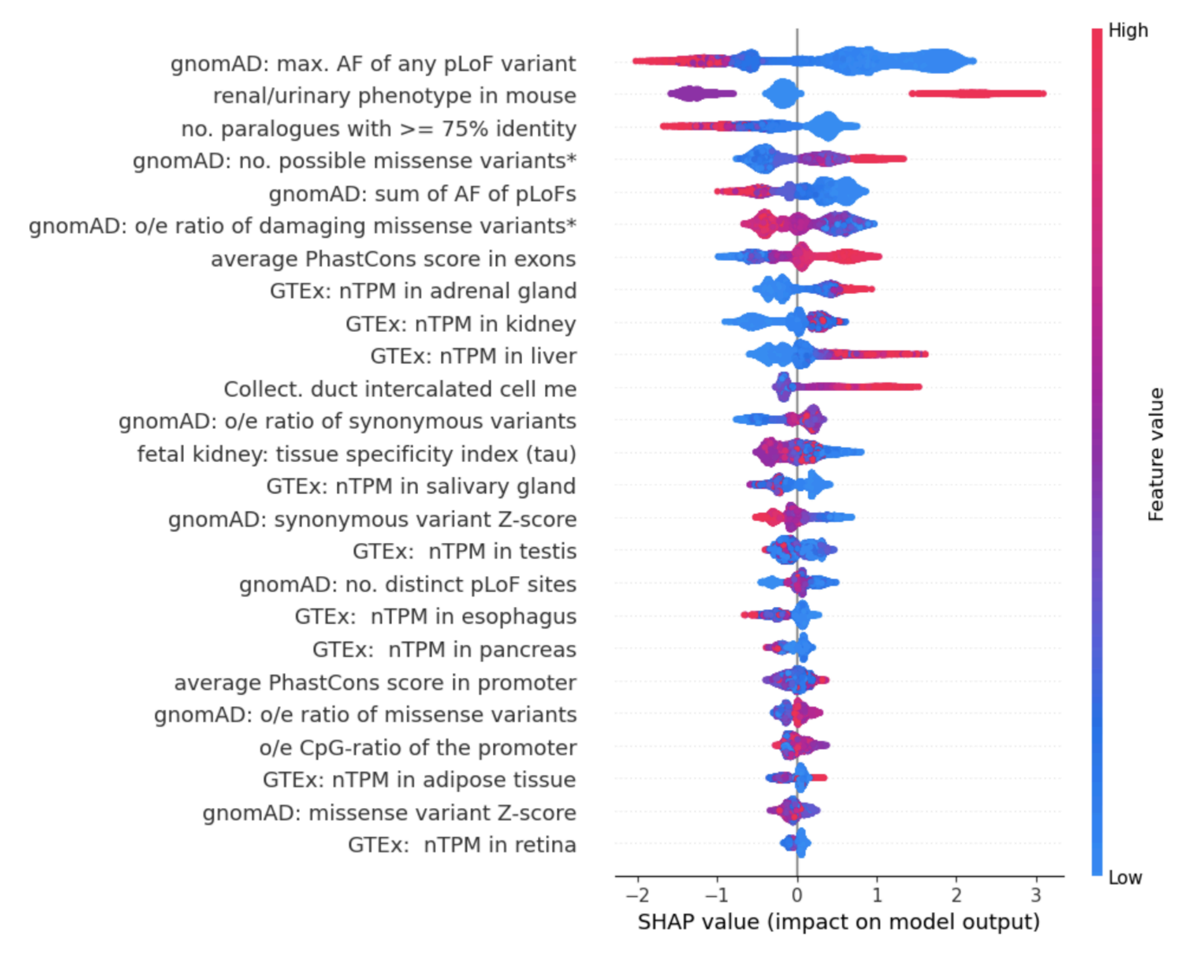


**Figure S1. SHAP beeswarm plot illustrating feature importance for the N-GS model.** Each point on the plot represents an individual gene. Features are ranked by their overall importance on the y-axis. For each feature, a point's position on the x-axis shows its impact on the model's raw output (expressed in log-odds as the SHAP values), while its color indicates the feature's original value (red for high, blue for low). Positive SHAP values push the log-odds of the N-GS prediction higher, increasing the likelihood of the gene being associated with kidney disease. me = ssRNA mean expression, pLoF = predicted-loss-of-function, AF = allele frequency, *predicted "probably damaging" by PolyPhen-2[38].

#### Section S9: Results of the GSEA performed on the N-GS predictions

The Gene Ontology (GO) terms GO:0003014 ("renal system process") and GO:0072001 ("renal system development"), including all their descendant terms, constituted a total of 633 terms. The 'GO_Biological_Process_2023' dataset in the Enrichr database contained 42 of these kidney terms and 5406 terms in total."

In the first analysis of our two-step GSEA, we assessed which specific gene sets were overrepresented among the top-ranked genes based on the N-GS. Fig. S2 depicts the top 5 enriched terms, while Table S4 summarizes the top-ranking GO-based gene sets along with their corresponding Enrichment Score (ES), Normalized Enrichment Score (NES), Nominal p-value (NOM p-val), False Discovery Rate (FDR q-val), Family-Wise Error Rate (FWER p-val) and Tag % values.

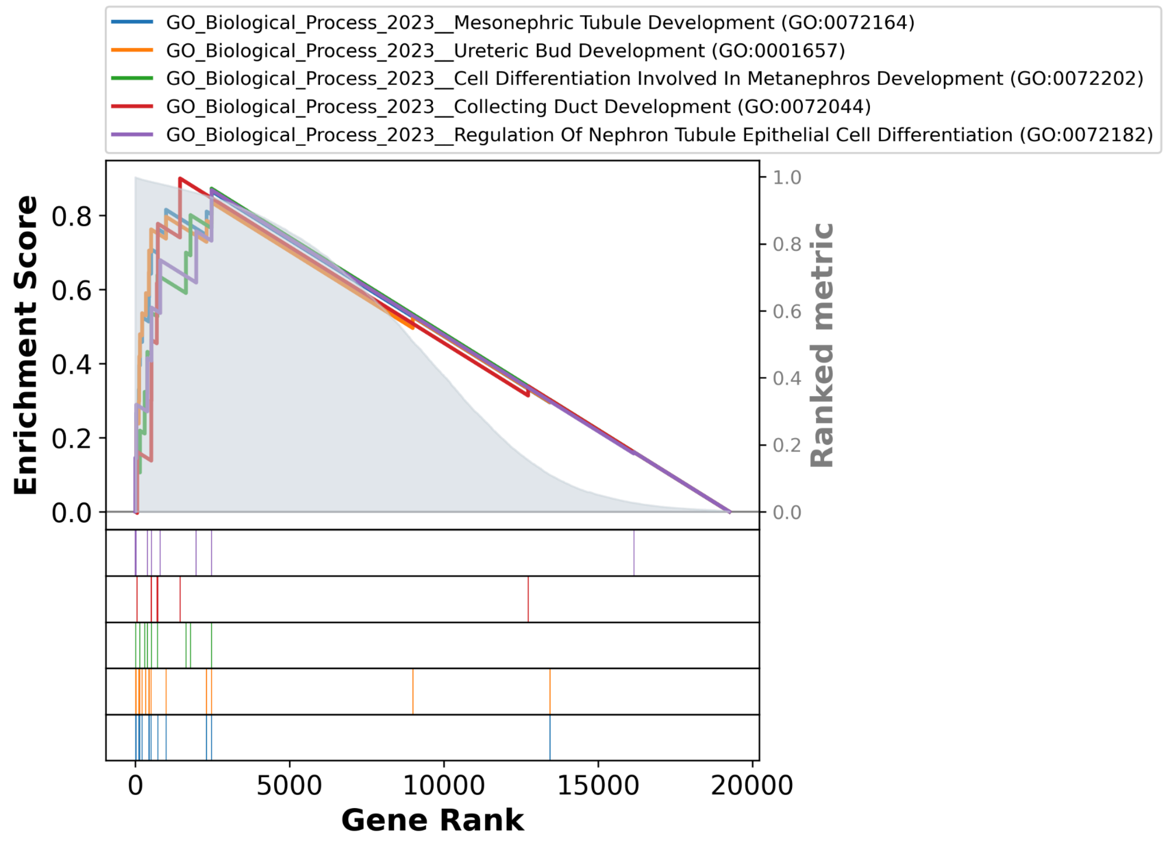


**Fig. S2. Enrichment plot of the top 5 enriched GO terms.** The analysis was performed on a list of all genes ranked by their Nephro Gene Score (N-GS), with the highest-scoring genes on the left. For each term, its corresponding curve shows the running Enrichment Score (ES), and the vertical black bars indicate the positions of genes from that set within the ranked list. The peak of the ES curve represents the point of maximum enrichment.

**Table S4. Top 20 Enriched GO Biological Processes Among Genes Ranked by Nephro Gene Score.** ES = Enrichment Score, NES = Normalized Enrichment Score, NOM p-val = Nominal p-value, FDR q-val = False Discovery Rate q-value, FWER p-val = Family-Wise Error Rate p-value.

| Process | ES | NES | NOM p-val | FDR q-val | FWER p-val | Tag % | kidney GO-term |
|---|---|---|---|---|---|---|---|
| Mesonephric Tubule Development (GO:0072164) | 0.865 | 1.942 | 0.0 | 0.0 | 0.0 | 15/16 | True |
| Ureteric Bud Development (GO:0001657) | 0.835 | 1.903 | 0.0 | 0.001 | 0.002 | 16/18 | True |
| Cell Differentiation Involved In Metanephros Development (GO:0072202) | 0.872 | 1.787 | 0.0 | 0.053 | 0.131 | 9/9 | True |
| Collecting Duct Development (GO:0072044) | 0.899 | 1.756 | 0.0 | 0.092 | 0.292 | 6/7 | True |
| Regulation Of Nephron Tubule Epithelial Cell Differentiation (GO:0072182) | 0.868 | 1.753 | 0.0 | 0.077 | 0.306 | 7/8 | True |
| Renal Filtration Cell Differentiation (GO:0061318) | 0.951 | 1.749 | 0.002 | 0.062 | 0.333 | 3/5 | True |
| Glomerular Epithelial Cell Differentiation (GO:0072311) | 0.951 | 1.749 | 0.002 | 0.062 | 0.333 | 3/5 | True |
| Metanephric Nephron Tubule Development (GO:0072234) | 0.925 | 1.748 | 0.0 | 0.055 | 0.339 | 6/6 | True |
| Regulation Of Peptidyl-Serine Phosphorylation Of STAT Protein (GO:0033139) | 0.776 | 1.738 | 0.0 | 0.065 | 0.415 | 2/18 | False |

| | | | | | | | |
|---|---|---|---|---|---|---|---|
| Regulation Of miRNA Metabolic Process (GO:2000628) | 0.873 | 1.735 | 0.0 | 0.064 | 0.437 | 8/8 | False |
| Regulation Of Transforming Growth Factor Beta2 Production (GO:0032909) | 0.913 | 1.726 | 0.0 | 0.07 | 0.497 | 5/6 | False |
| Positive Regulation Of Peptidyl-Serine Phosphorylation Of STAT Protein (GO:0033141) | 0.778 | 1.726 | 0.0 | 0.066 | 0.501 | 2/17 | False |
| Podocyte Differentiation (GO:0072112) | 0.804 | 1.701 | 0.001 | 0.104 | 0.698 | 8/11 | True |
| Biomineral Tissue Development (GO:0031214) | 0.838 | 1.698 | 0.001 | 0.102 | 0.716 | 4/9 | False |
| Metanephros Development (GO:0001656) | 0.728 | 1.697 | 0.0 | 0.098 | 0.723 | 20/25 | True |
| Regulation Of Keratinocyte Apoptotic Process (GO:1902172) | 0.93 | 1.696 | 0.0 | 0.094 | 0.732 | 3/5 | False |
| Epithelial Tube Formation (GO:0072175) | 0.929 | 1.693 | 0.0 | 0.093 | 0.76 | 5/5 | False |
| Metanephric Collecting Duct Development (GO:0072205) | 0.925 | 1.687 | 0.0 | 0.099 | 0.794 | 5/5 | True |
| CRD-mediated mRNA Stabilization (GO:0070934) | 0.804 | 1.675 | 0.001 | 0.119 | 0.866 | 9/10 | False |
| Galactosylceramide Metabolic Process (GO:0006681) | 0.853 | 1.668 | 0.0 | 0.127 | 0.896 | 5/7 | False |

#### Section S10: Model Performance of the N-VS

Table S5 presents the performance metrics of the N-VS in both the training and the test set.

**Table S5. Performance metrics of the Nephro Variant Score across the training and test set**. The threshold dependent metrics were yielded based on a threshold of 0.5 for positive/negative classification.

| | Training set | Test set |
|---|---|---|
| **Area-under-curve (AUC)** | 0.991 | 0.994 |
| **Precsion-Recall-AUC** | 0.984 | 0.986 |
| **Accuracy** | 0.955 | 0.974 |
| **Sensitivity** | 0.957 | 0.963 |
| **Specificity** | 0.954 | 0.978 |
| **False-discovery-rate** | 0.08 | 0.064 |
| **False-positive-rate** | 0.046 | 0.022 |
| **Negative predictive value** | 0.976 | 0.987 |
| **Positive predictive value** | 0.92 | 0.936 |

Additionally, we evaluated the AUC across different functional impact groups, with the results presented in Table S6. Note that the overall AUC is higher than the individual AUCs for each impact subgroup. Since a variant's predicted functional impact was the most influential predictor, the within-subset discrimination task was more challenging than the overall task, as the model must distinguish among more homogeneous subsets with reduced variation in this key feature.

| Table S6. Area-under-curve (AUC) of the Nephro Variant Score predictions across different variant impact groups. | | | | |
|---|---|---|---|---|
| **Impact** | MODIFIER | LOW | MODERATE | HIGH |
| **AUC** | 0.857 | 0.965 | 0.933 | 0.847 |

#### Section S11: Clinical description of patients carrying high-scoring *ERMP1* variants

| **Table S7: Clinical description and genetic findings of patients carrying high-scoring ERMP1 variants.** |
|---|
| Patient 1 (N-CS = 8.0): male in his 60s, suspected Alport syndrome (based on histology), multiple kidney cysts, ESRD, likely pathogenic variant in COL4A3 and VUS in COL4A4 |
| Patient 2 (N-CS = 8.0): male in his 50s, suspected Alport syndrome (histology: thin basement membrane nephropathy and mild hypertensive nephropathy), diabetes mellitus type 2, CKDG3aA3, no diagnostic variants found |
| Patient 3 (N-CS = 8.0): female in her 40s, ADPKD-PKD1, Mayo 1A-B, CKDG2A1, pathogenic variant in PKD1 |
| Patient 4: (N-CS = 7.8): male in his 20s, suspected autosomal-dominant Alport syndrome (microhematuria), eGFR normal, pathogenic variant in COL4A4 |

#### Section S12: Local SHAP Feature Contribution Analysis for *ERMP1*

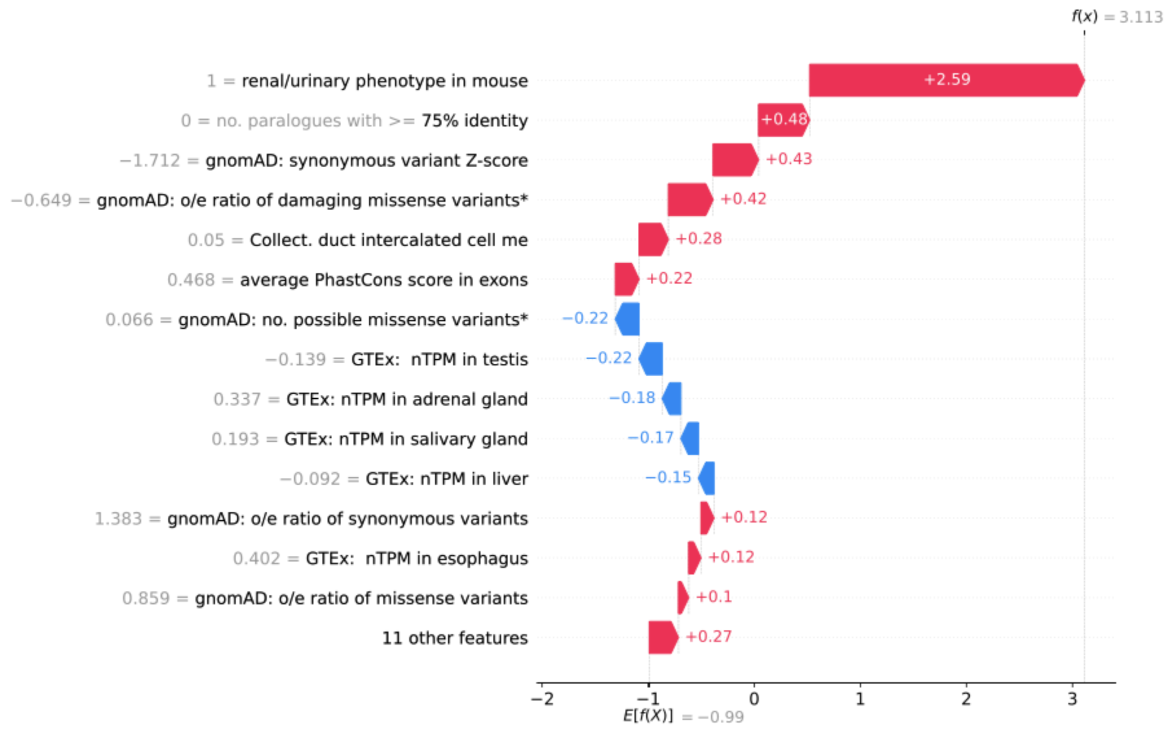


**Fig. S3. SHAP waterfall plot illustrating the local feature contributions for the gene *ERMP1*.** The starting point on the x-axis (E[f(X)] = -0.99) represents the model's baseline expected value (the average log-odds prediction across the entire training dataset). The final value at the top (f(x) = 3.113) represents the model's final predicted log-odds score for *ERMP1*. Features are listed on the y-axis in descending order of their absolute impact on this specific prediction. The light grey numbers on the left indicate the specific input values for *ERMP1* (either raw or scaled, depending on model preprocessing). Red arrows denote features that push the log-odds higher (increasing the likelihood of the gene being associated with kidney disease), while blue arrows denote features that decrease the log-odds. me = ssRNA mean expression, pLoF = predicted-loss-of-function, AF = allele frequency, *predicted "probably damaging" by PolyPhen-2 [38].

#### Section S13: Evaluation of Exome-Wide Gene Burden Counts Omitting the N-CS Filter

To evaluate the specific impact of the N-CS framework on prioritizing high-confidence disease drivers, we performed a comparative baseline analysis by setting the minimum score threshold to 0. All other downstream quality control parameters, technical region annotations were maintained as described in Section S5.

Omitting the N-CS filter resulted in a massive expansion of candidate genes meeting the event threshold across the cohort (Fig. S4). While this unfiltered analysis successfully captures known drivers, the top signals are heavily populated by a broad pool of unprioritized candidates and genes of uncertain clinical or renal significance, demonstrating the necessity of the N-CS threshold in isolating true renal disease candidates from background genetic variance.

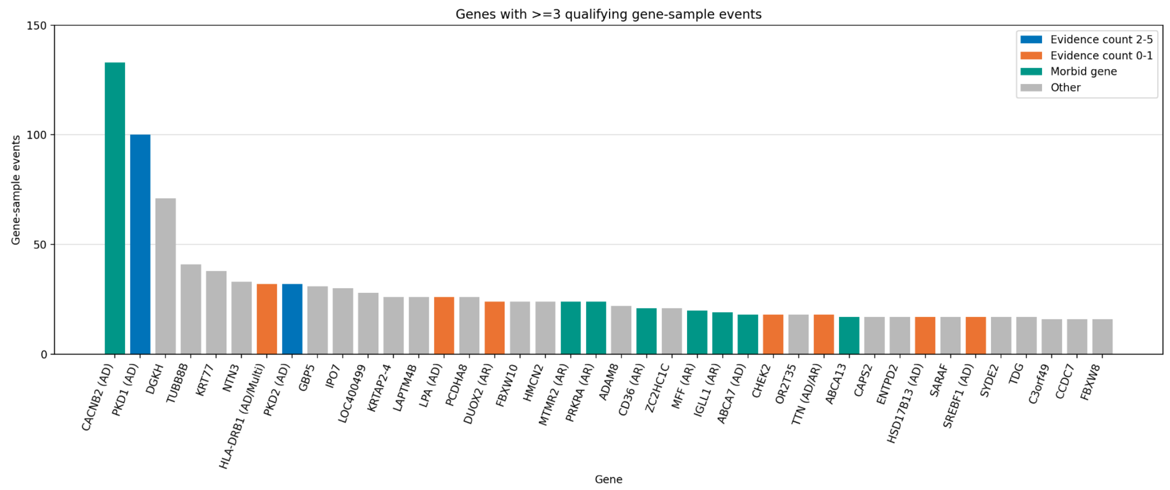


**Fig. S4. Exome-Wide Gene Burden Analysis Without N-CS Prioritization.** Bar plot displaying candidate genes accumulating ≥3 qualifying independent gene-sample events when the N-CS filter is omitted from the pipeline. AD = autosomal dominant, AR = autosomal recessive. Note the immense increase in the number of candidate genes and total event counts compared to the prioritized main figure.

#### Section S14: PKD Case-Control Association and Rank Shift Analysis

The magnitude and direction of the associations for genes achieving nominal, unadjusted statistical significance ($p \leq 0.05$) are visualized using continuity-corrected $\log_2$-odds ratios (Fig. S5).

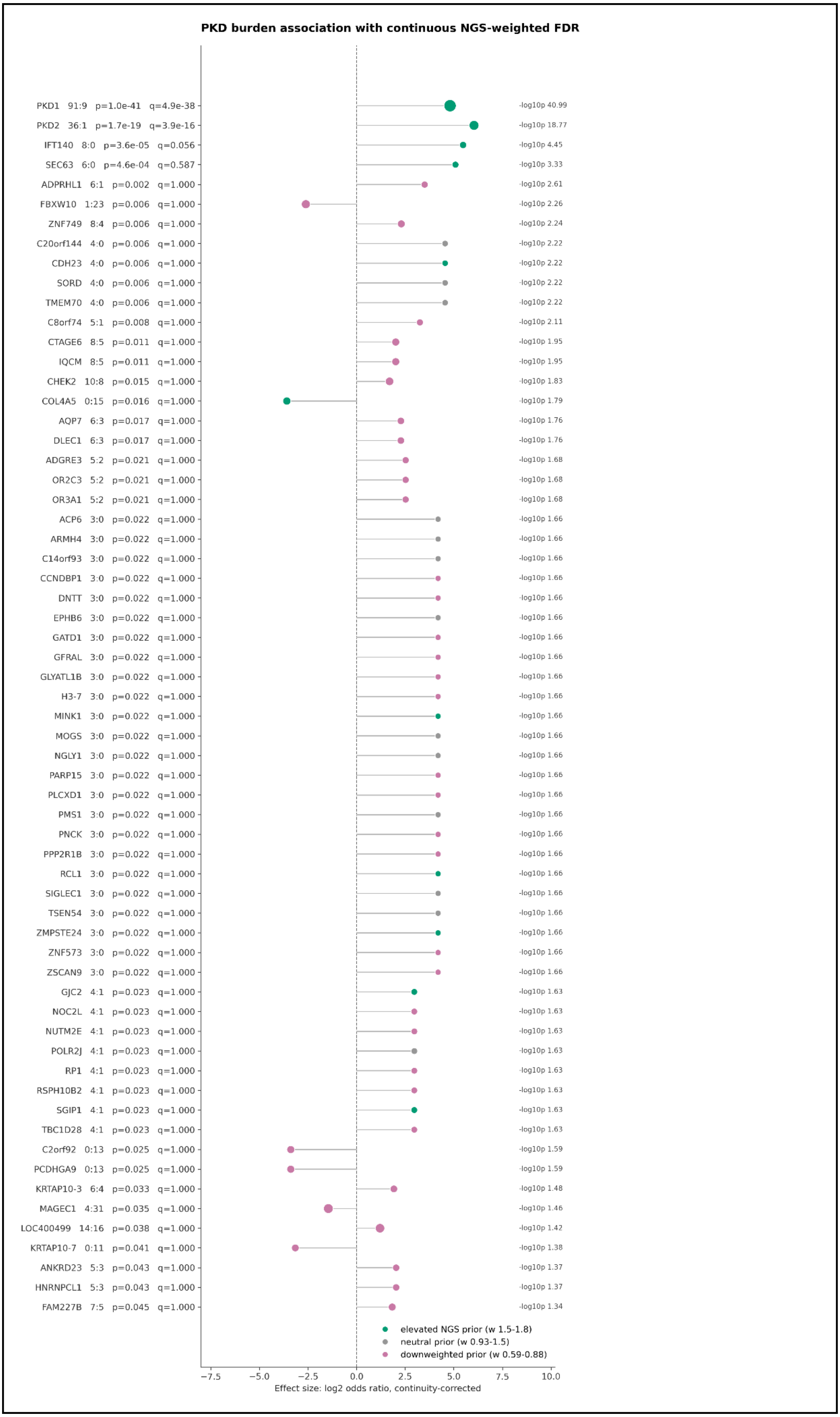
PKD burden association with continuous NGS-weighted FDR
PKD1 91:9 p=1.0e-41 q=4.9e-38
-log10p 40.99
PKD2 36:1 p=1.7e-19 q=3.9e-16
-log10p 18.77
IFT140 8:0 p=3.6e-05 q=0.056
-log10p 4.45
SEC63 6:0 p=4.6e-04 q=0.587
-log10p 3.33
ADPRHL1 6:1 p=0.002 q=1.000
-log10p 2.61
FBXW10 1:23 p=0.006 q=1.000
-log10p 2.26
ZNF749 8:4 p=0.006 q=1.000
-log10p 2.24
C20orf144 4:0 p=0.006 q=1.000
-log10p 2.22
CDH23 4:0 p=0.006 q=1.000
-log10p 2.22
SORD 4:0 p=0.006 q=1.000
-log10p 2.22
TMEM70 4:0 p=0.006 q=1.000
-log10p 2.22
C8orf74 5:1 p=0.008 q=1.000
-log10p 2.11
CTAGE6 8:5 p=0.011 q=1.000
-log10p 1.95
IQCM 8:5 p=0.011 q=1.000
-log10p 1.95
CHEK2 10:8 p=0.015 q=1.000
-log10p 1.83
COL4A5 0:15 p=0.016 q=1.000
-log10p 1.79
AQP7 6:3 p=0.017 q=1.000
-log10p 1.76
DLEC1 6:3 p=0.017 q=1.000
-log10p 1.76
ADGRE3 5:2 p=0.021 q=1.000
-log10p 1.68
OR2C3 5:2 p=0.021 q=1.000
-log10p 1.68
OR3A1 5:2 p=0.021 q=1.000
-log10p 1.68
ACP6 3:0 p=0.022 q=1.000
-log10p 1.66
ARMH4 3:0 p=0.022 q=1.000
-log10p 1.66
C14orf93 3:0 p=0.022 q=1.000
-log10p 1.66
CCNDBP1 3:0 p=0.022 q=1.000
-log10p 1.66
DNTT 3:0 p=0.022 q=1.000
-log10p 1.66
EPHB6 3:0 p=0.022 q=1.000
-log10p 1.66
GATD1 3:0 p=0.022 q=1.000
-log10p 1.66
GFRAL 3:0 p=0.022 q=1.000
-log10p 1.66
GLYATL1B 3:0 p=0.022 q=1.000
-log10p 1.66
H3-7 3:0 p=0.022 q=1.000
-log10p 1.66
MINK1 3:0 p=0.022 q=1.000
-log10p 1.66
MOGS 3:0 p=0.022 q=1.000
-log10p 1.66
NGLY1 3:0 p=0.022 q=1.000
-log10p 1.66
PARP15 3:0 p=0.022 q=1.000
-log10p 1.66
PLCXD1 3:0 p=0.022 q=1.000
-log10p 1.66
PMS1 3:0 p=0.022 q=1.000
-log10p 1.66
PNCK 3:0 p=0.022 q=1.000
-log10p 1.66
PPP2R1B 3:0 p=0.022 q=1.000
-log10p 1.66
RCL1 3:0 p=0.022 q=1.000
-log10p 1.66
SIGLEC1 3:0 p=0.022 q=1.000
-log10p 1.66
TSEN54 3:0 p=0.022 q=1.000
-log10p 1.66
ZMPSTE24 3:0 p=0.022 q=1.000
-log10p 1.66
ZNF573 3:0 p=0.022 q=1.000
-log10p 1.66
ZSCAN9 3:0 p=0.022 q=1.000
-log10p 1.66
GJC2 4:1 p=0.023 q=1.000
-log10p 1.63
NOC2L 4:1 p=0.023 q=1.000
-log10p 1.63
NUTM2E 4:1 p=0.023 q=1.000
-log10p 1.63
POLR2J 4:1 p=0.023 q=1.000
-log10p 1.63
RP1 4:1 p=0.023 q=1.000
-log10p 1.63
RSPH10B2 4:1 p=0.023 q=1.000
-log10p 1.63
SGIP1 4:1 p=0.023 q=1.000
-log10p 1.63
TBC1D28 4:1 p=0.023 q=1.000
-log10p 1.63
C2orf92 0:13 p=0.025 q=1.000
-log10p 1.59
PCDHGA9 0:13 p=0.025 q=1.000
-log10p 1.59
KRTAP10-3 6:4 p=0.033 q=1.000
-log10p 1.48
MAGEC1 4:31 p=0.035 q=1.000
-log10p 1.46
LOC400499 14:16 p=0.038 q=1.000
-log10p 1.42
KRTAP10-7 0:11 p=0.041 q=1.000
-log10p 1.38
ANKRD23 5:3 p=0.043 q=1.000
-log10p 1.37
HNRNPCL1 5:3 p=0.043 q=1.000
-log10p 1.37
FAM227B 7:5 p=0.045 q=1.000
-log10p 1.34
elevated NGS prior (w 1.5-1.8)
neutral prior (w 0.93-1.5)
downweighted prior (w 0.59-0.88)
−7.5
−5.0
−2.5
0.0
2.5
5.0
7.5
10.0
Effect size: log2 odds ratio, continuity-corrected

**Fig. S5. Effect sizes and cohort metrics for nominally significant genes in the PKD association analysis.** The plot displays genes reaching nominal significance $p \leq 0.05$ prior to multiple-testing correction. The x-axis represents the continuity-corrected $log_2$-odds ratio. Genes on the y-axis are ordered by their adjusted weighted significance ($q_w$). Marker size is proportional to the total number of qualifying variant alleles. Target genes of high biological interest (*PKD1*, *PKD2*, *IFT140*, and *SEC63*) are highlighted in orange. For the remaining background genes, marker colors indicate the N-GS weighting tier applied during FDR correction: green denotes an elevated prior (weight >= 1.5), grey denotes a neutral prior (0.9 <= weight < 1.5), and pink denotes a downweighted prior (weight <= 0.9).

The rank shift evaluation visualizes the statistical impact of the N-GS biological prior within the weighted False Discovery Rate (FDR) framework (Fig. S6). The integration of the prior systematically redistributed the statistical penalty across the nominally significant genes. Candidates with low N-GS values received stricter penalties (downweighted priors), shifting their adjusted significance leftward, further away from the exome-wide threshold. Conversely, established genes with high N-GS values received a relaxed penalty. While the massive effect sizes of the primary drivers *PKD1* and *PKD2* rendered them exome-wide significant regardless of the correction method, the weighting framework actively enhanced the statistical prioritization of secondary candidates like *IFT140*, driving a rightward shift that brought it to the cusp of the formal significance threshold ($q_w = 0.056$).

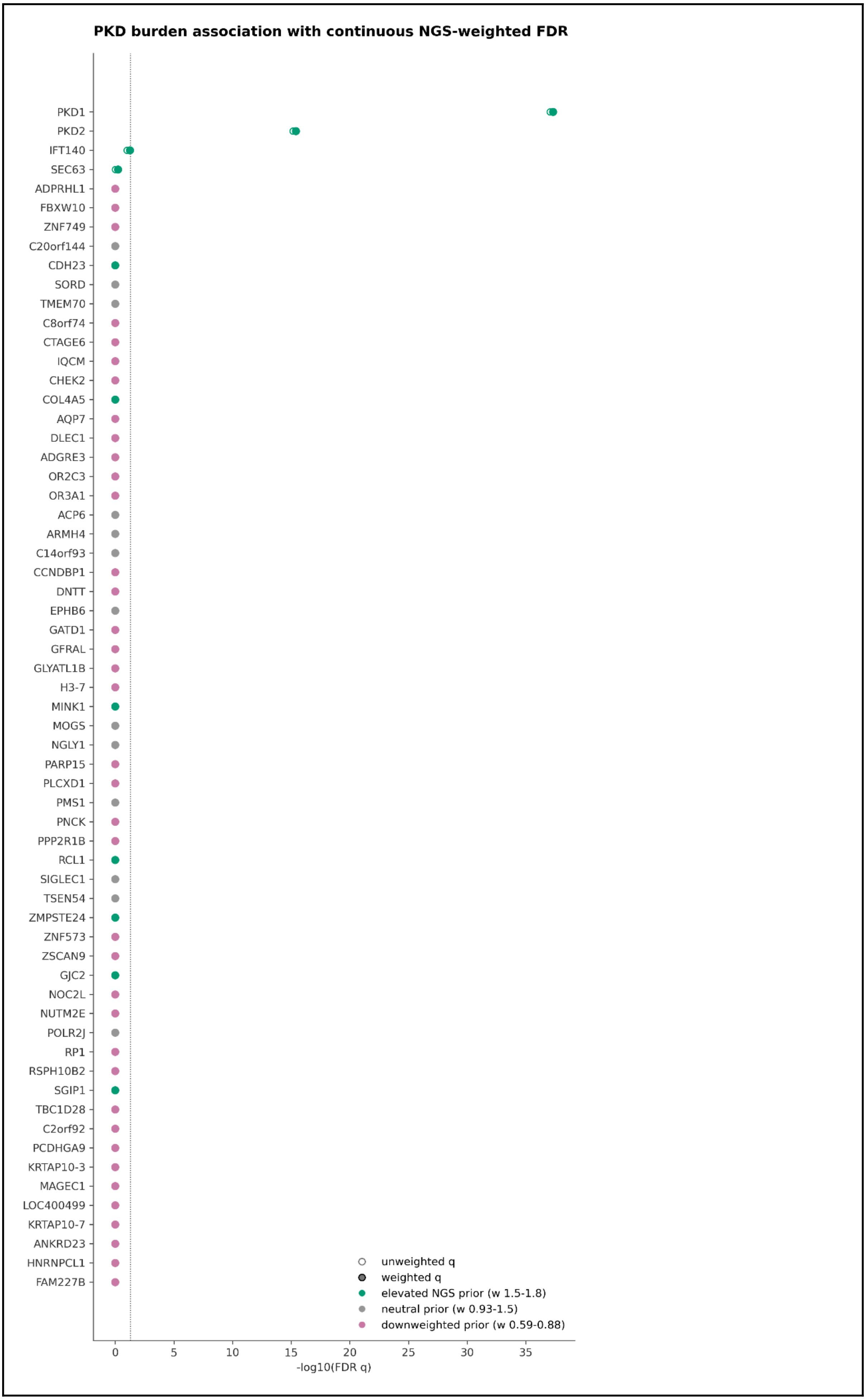
PKD burden association with continuous NGS-weighted FDR
PKD1
PKD2
IFT140
SEC63
ADPRHL1
FBXW10
ZNF749
C20orf144
CDH23
SORD
TMEM70
C8orf74
CTAGE6
IQCM
CHEK2
COL4A5
AQP7
DLEC1
ADGRE3
OR2C3
OR3A1
ACP6
ARMH4
C14orf93
CCNDBP1
DNTT
EPHB6
GATD1
GFRAL
GLYATL1B
H3-7
MINK1
MOGS
NGLY1
PARP15
PLCXD1
PMS1
PNCK
PPP2R1B
RCL1
SIGLEC1
TSEN54
ZMPSTE24
ZNF573
ZSCAN9
GJC2
NOC2L
NUTM2E
POLR2J
RP1
RSPH10B2
SGIP1
TBC1D28
C2orf92
PCDHGA9
KRTAP10-3
MAGEC1
LOC400499
KRTAP10-7
ANKRD23
HNRNPCL1
FAM227B
unweighted q
weighted q
elevated NGS prior (w 1.5-1.8)
neutral prior (w 0.93-1.5)
downweighted prior (w 0.59-0.88)
0
5
10
15
20
25
30
35
-log10(FDR q)

**Fig. S6. Rank shift evaluation demonstrating the impact of the N-GS biological prior.** The plot visualizes the shift in adjusted statistical significance for nominally associated genes when transitioning from a standard, unweighted correction to the continuous N-GS-weighted False Discovery Rate (FDR) framework. Highlighted genes of interest are shown in orange. A rightward shift indicates a relaxed penalty for biologically plausible candidates (high N-GS), whereas a leftward shift indicates a stricter penalty for genes lacking established renal evidence (low N-GS).

#### Section S15: Variants and their N-CS of Novel Candidate Genes

Table S8 lists the 46 variants from the 11 novel candidate genes identified in the literature search together with their mode of inheritance, N-GS, N-VS, IS and the composite N-CS.

**Table S8. Variants and N-CS of novel candidate genes.**

| Variant | Gene | Inheritance | Disease | Ref | Journal | N-GS | N-VS | IS | N-CS |
|---|---|---|---|---|---|---|---|---|---|
| chr19-926060-G-C | ARID3A | dominant | CAKUT | 39 | Nat Commun | 0.91 | 1.0 | 0.32 | 8.28 |
| chr19-929604-C-T | ARID3A | unknown | CAKUT | 39 | Nat Commun | 0.91 | 1.0 | 0.1 | 7.84 |
| chr19-929838-C-T | ARID3A | de novo | CAKUT | 39 | Nat Commun | 0.91 | 1.0 | 0.95 | 9.54 |
| chr19-964432-G-C | ARID3A | unknown | CAKUT | 39 | Nat Commun | 0.91 | 1.0 | 0.1 | 7.84 |
| chr19-964831-A-T | ARID3A | unknown | CAKUT | 39 | Nat Commun | 0.91 | 1.0 | 0.1 | 7.84 |
| chr19-964918-C-T | ARID3A | unknown | CAKUT | 39 | Nat Commun | 0.91 | 1.0 | 0.1 | 7.84 |
| chr19-966631-CACCCT-C | ARID3A | de novo | CAKUT | 39 | Nat Commun | 0.91 | 0.99 | 0.95 | 9.52 |
| chr9-124554439-G-A | NR6A1 | dominant | CAKUT | 39 | Nat Commun | 0.85 | 0.95 | 0.32 | 7.82 |
| chr9-124554439-G-A | NR6A1 | dominant | CAKUT | 39 | Nat Commun | 0.85 | 0.95 | 0.32 | 7.82 |
| chr9-124554439-G-A | NR6A1 | dominant | CAKUT | 40 | Nat Commun | 0.85 | 0.95 | 0.32 | 7.82 |
| chr9-124554516-C-T | NR6A1 | unknown | CAKUT | 39 | Nat Commun | 0.85 | 0.92 | 0.1 | 7.29 |
| chr9-124554367-G-A | NR6A1 | unknown | CAKUT | 39 | Nat Commun | 0.85 | 0.95 | 0.1 | 7.38 |
| chr9-124524769-G-A | NR6A1 | unknown | CAKUT | 40 | Nat Commun | 0.85 | 0.95 | 0.1 | 7.38 |
| chr9-124535976-AGCTCCTGCCACGTAG-T | NR6A1 | dominant | CAKUT | 40 | Nat Commun | 0.85 | 0.37 | 0.32 | 5.51 |

| | | | | | | | | | |
|---|---|---|---|---|---|---|---|---|---|
| chr9-124536055-C-T | NR6A1 | unknown | CAKUT | 40 | Nat Commun | 0.85 | 1.0 | 0.1 | 7.59 |
| chr9-124554424-T-TG | NR6A1 | de novo | CAKUT | 40 | Nat Commun | 0.85 | 0.99 | 0.95 | 9.26 |
| chr1-204123131-C-A | SOX13 | de novo | CAKUT | 41 | Genet Med | 0.93 | 1.0 | 0.95 | 9.61 |
| chr6-78990940-C-CT | PHIP | de novo | CAKUT | 42 | Eur J Hum Genet | 0.99 | 0.99 | 0.95 | 9.83 |
| chr6-79015155-G-A | PHIP | de novo | CAKUT | 42 | Eur J Hum Genet | 0.99 | 0.94 | 0.95 | 9.6 |
| chr6-79078051-T-G | PHIP | unknown | CAKUT | 42 | Eur J Hum Genet | 0.99 | 0.61 | 0.1 | 6.58 |
| chr6-78958572-G-A | PHIP | unknown | CAKUT | 42 | Eur J Hum Genet | 0.99 | 1.0 | 0.1 | 8.16 |
| chr6-78946136-C-T | PHIP | unknown | CAKUT | 42 | Eur J Hum Genet | 0.99 | 0.7 | 0.1 | 6.96 |
| chr2-128108039-TACTC-T | UGGT1 | comp. het. | Cystic, Syndromic | 43 | Am J Hum Genet | 0.98 | 1.0 | 0.64 | 9.18 |
| chr2-128127389-CTGGAGATTCAGCCCTCTTCATCAA-C | UGGT1 | comp. het. | Cystic, Syndromic | 43 | Am J Hum Genet | 0.98 | 0.37 | 0.64 | 6.66 |
| chr2-128121203-TCA-T | UGGT1 | hom. recessive | Cystic, Syndromic | 43 | Am J Hum Genet | 0.98 | 0.99 | 0.64 | 9.15 |
| chr2-128187608-C-T | UGGT1 | hom. recessive | Cystic, Syndromic | 43 | Am J Hum Genet | 0.98 | 1.0 | 0.64 | 9.18 |
| chr2-128187608-C-T | UGGT1 | hom. recessive | Cystic, Syndromic | 43 | Am J Hum Genet | 0.98 | 1.0 | 0.64 | 9.18 |
| chr2-128187608-C-T | UGGT1 | hom. recessive | Cystic, Syndromic | 43 | Am J Hum Genet | 0.98 | 1.0 | 0.64 | 9.18 |
| chr2-128187608-C-T | UGGT1 | hom. recessive | Cystic, Syndromic | 43 | Am J Hum Genet | 0.98 | 1.0 | 0.64 | 9.18 |
| chr2-128152899-C-T | UGGT1 | hom. recessive | Cystic, Syndromic | 43 | Am J Hum Genet | 0.98 | 0.94 | 0.64 | 8.94 |
| chrX-118442587-C-T | WDR44 | X-linked recessive | Ciliopathy, Syndromic | 44 | Nat Commun | 0.93 | 0.85 | 0.56 | 8.25 |

| | | | | | | | | | |
|---|---|---|---|---|---|---|---|---|---|
| chrX-118442640-G-T | WDR44 | X-linked recessive | Ciliopathy, Syndromic | [44] | Nat Commun | 0.93 | 0.86 | 0.56 | 8.28 |
| chrX-118442274-C-T | WDR44 | X-linked recessive | Ciliopathy, Syndromic | [44] | Nat Commun | 0.93 | 1.0 | 0.56 | 8.84 |
| chrX-118436793-A-G | WDR44 | X-linked recessive | Ciliopathy, Syndromic | [44] | Nat Commun | 0.93 | 0.83 | 0.56 | 8.15 |
| chrX-118441396-T-C | WDR44 | X-linked recessive | Ciliopathy, Syndromic | [44] | Nat Commun | 0.93 | 0.81 | 0.56 | 8.06 |
| chrX-118441396-T-C | WDR44 | X-linked recessive | Ciliopathy, Syndromic | [44] | Nat Commun | 0.93 | 0.81 | 0.56 | 8.06 |
| chrX-118441398-G-A | WDR44 | X-linked recessive | Ciliopathy, Syndromic | [44] | Nat Commun | 0.93 | 0.92 | 0.56 | 8.52 |
| chrX-118441549-A-G | WDR44 | X-linked recessive | Ciliopathy, Syndromic | [44] | Nat Commun | 0.93 | 0.84 | 0.56 | 8.2 |
| chrX-118444366-A-G | WDR44 | X-linked recessive | Ciliopathy, Syndromic | [44] | Nat Commun | 0.93 | 0.63 | 0.56 | 7.37 |
| chr2-121231887-TG-T | TFCP2L1 | hom. recessive | Salt wasting | [45] | Pediatr Nephrol | 1.0 | 0.99 | 0.64 | 9.24 |
| chr16-88738318-C-T | PIEZO1 | comp. het. | CAKUT, Syndromic | [46] | Nat Commun | 0.98 | 0.33 | 0.64 | 6.51 |
| chr16-88717099-G-A | PIEZO1 | comp. het. | CAKUT, Syndromic | [46] | Nat Commun | 0.98 | 0.85 | 0.64 | 8.61 |
| chr19-31279621-T-C | TSHZ3 | dominant | CAKUT, Syndromic | [47] | Eur J Hum Genet | 0.99 | 0.01 | 0.32 | 4.64 |
| chr19-31279621-T-C | TSHZ3 | dominant | CAKUT, Syndromic | [47] | Eur J Hum Genet | 0.99 | 0.01 | 0.32 | 4.64 |
| chr19-31279621-T-C | TSHZ3 | dominant | CAKUT, Syndromic | [47] | Eur J Hum Genet | 0.99 | 0.01 | 0.32 | 4.64 |
| chr19-31279621-T-C | TSHZ3 | dominant | CAKUT, Syndromic | [47] | Eur J Hum Genet | 0.99 | 0.01 | 0.32 | 4.64 |
| chr19-31279621-T-C | TSHZ3 | dominant | CAKUT, Syndromic | [47] | Eur J Hum Genet | 0.99 | 0.01 | 0.32 | 4.64 |
| chr19-31279605-G-A | TSHZ3 | dominant | CAKUT, Syndromic | [47] | Eur J Hum Genet | 0.99 | 0.69 | 0.32 | 7.35 |

| | | | | | | | | | |
|---|---|---|---|---|---|---|---|---|---|
| chr19-31279597-C-T | TSHZ3 | de novo | CAKUT, Syndromic | [47] | Eur J Hum Genet | 0.99 | 0.56 | 0.95 | 8.13 |
| chr19-31277914-T-C | TSHZ3 | dominant | CAKUT, Syndromic | [47] | Eur J Hum Genet | 0.99 | 0.56 | 0.32 | 6.87 |
| chr19-31279564-G-A | TSHZ3 | dominant | CAKUT, Syndromic | [47] | Eur J Hum Genet | 0.99 | 0.56 | 0.32 | 6.86 |
| chrX-35975809-G-A | CFAP47 | X-linked recessive | Cystic | [48] | Kidney Int Rep | 0.03 | 0.02 | 0.56 | 1.34 |
| chrX-35966701-T-G | CFAP47 | X-linked recessive | Cystic | [48] | Kidney Int Rep | 0.03 | 0.56 | 0.56 | 3.51 |
| chrX-35919816-G-A | CFAP47 | X-linked recessive | Cystic | [48] | Kidney Int Rep | 0.03 | 0.04 | 0.56 | 1.4 |
| chr17-1483665-G-A | MYO1C | hom. recessive | Nephrotic | [49] | Pediatr Nephrol | 0.87 | 0.75 | 0.64 | 7.75 |
| chr17-1470629-T-A | MYO1C | hom. recessive | Nephrotic | [49] | Pediatr Nephrol | 0.87 | 0.59 | 0.64 | 7.13 |

#### References


1. Pedregosa F, Varoquaux G, Gramfort A, et al. Scikit-learn: Machine Learning in Python. *J Mach Learn Res*. 2011;12(Oct):2825-2830.

2. HCA seed network precise tumor-nephrectomy samples. Accessed August 25, 2025. https://explore.data.humancellatlas.org/projects/29ed827b-c539-4f4c-bb6b-ce8f9173dfb7

3. Cellxgene Data Portal. Cellxgene Data Portal. Accessed August 25, 2025. https://cellxgene.cziscience.com/collections/a98b828a-622a-483a-80e0-15703678befd

4. Cao J, O'Day DR, Pliner HA, et al. A human cell atlas of fetal gene expression. *Science*. 2020;370(6518):eaba7721.

5. Yanai I, Benjamin H, Shmoish M, et al. Genome-wide midrange transcription profiles reveal expression level relationships in human tissue specification. *Bioinformatics*. 2005;21(5):650-659.

6. Karczewski KJ, Francioli LC, Tiao G, et al. The mutational constraint spectrum quantified from variation in 141,456 humans. *Nature*. 2020;581(7809):434-443.

7. The Human Protein Atlas. Accessed August 25, 2025. https://v23.proteinatlas.org/

8. Cunningham F, Allen JE, Allen J, et al. Ensembl 2022. *Nucleic Acids Res*. 2022;50(D1):D988-D995.

9. MGI-Mouse Genome Informatics-The international database resource for the laboratory mouse. Accessed August 25, 2025. http://www.informatics.jax.org)

10. Subramanian A, Tamayo P, Mootha VK, et al. Gene set enrichment analysis: a knowledge-based approach for interpreting genome-wide expression profiles. *Proc Natl Acad Sci U S A*. 2005;102(43):15545-15550.

11. Fang Z, Liu X, Peltz G. GSEApy: a comprehensive package for performing gene set enrichment analysis in Python. *Bioinformatics*. 2023;39(1). doi:10.1093/bioinformatics/btac757

12. Xie Z, Bailey A, Kuleshov MV, et al. Gene set knowledge discovery with Enrichr. *Curr Protoc*. 2021;1(3):e90.

13. McLaren W, Gil L, Hunt SE, et al. The Ensembl Variant Effect Predictor. *Genome Biol*. 2016;17(1):122.

14. Harrison PW, Amode MR, Austine-Orimoloye O, et al. Ensembl 2024. *Nucleic Acids Res*. 2024;52(D1):D891-D899.

15. Lundberg S, Lee SI. A unified approach to interpreting model predictions. *arXiv [csAI]*. Published online 2017. doi:10.48550/ARXIV.1705.07874

16. McLaren W, Pritchard B, Rios D, Chen Y, Flicek P, Cunningham F. Deriving the consequences of genomic variants with the Ensembl API and SNP Effect Predictor. *Bioinformatics*. 2010;26(16):2069-2070.

17. Rentzsch P, Witten D, Cooper GM, Shendure J, Kircher M. CADD: predicting the deleteriousness of variants throughout the human genome. *Nucleic Acids Res*. 2019;47(D1):D886-D894.

18. Jaganathan K, Kyriazopoulou Panagiotopoulou S, McRae JF, et al. Predicting splicing from primary sequence with deep learning. *Cell*. 2019;176(3):535-548.e24.

19. Siepel A, Bejerano G, Pedersen JS, et al. Evolutionarily conserved elements in vertebrate, insect, worm, and yeast genomes. *Genome Res*. 2005;15(8):1034-1050.

20. McKenna A, Hanna M, Banks E, et al. The Genome Analysis Toolkit: a MapReduce framework for analyzing next-generation DNA sequencing data. *Genome Res*. 2010;20(9):1297-1303.

21. Popp B. *Varvis-Download*. Github. Accessed September 22, 2026. https://github.com/LaborBerlin/varvis-download

22. Popp B. *Hardnormly*. Github. Accessed May 29, 2026. https://github.com/berntpopp/hardnormly

23. Popp B. *Merge-Multisample-Vcf*. Github. Accessed September 22, 2026. https://github.com/scholl-lab/merge-multisample-vcf

24. Danecek P, Bonfield JK, Liddle J, et al. Twelve years of SAMtools and BCFtools. *Gigascience*. 2021;10(2). doi:10.1093/gigascience/giab008

25. Popp B. *Variantcentrifuge: A Pipeline for Filtering Annotated Variant Call Format Files*. Github. Accessed May 29, 2026. https://github.com/scholl-lab/variantcentrifuge

26. Mölder F, Jablonski KP, Letcher B, et al. Sustainable data analysis with Snakemake. *F1000Res*. 2021;10:33.

27. Popp B. *Sm-Vcf-Annotation*. Github. Accessed May 29, 2026. https://github.com/scholl-lab/sm-vcf-annotation

28. Morales J, Pujar S, Loveland JE, et al. A joint NCBI and EMBL-EBI transcript set for clinical genomics and research. *Nature*. 2022;604(7905):310-315.

29. Cingolani P, Platts A, Wang LL, et al. A program for annotating and predicting the effects of single nucleotide polymorphisms, SnpEff: SNPs in the genome of Drosophila melanogaster strain w1118; iso-2; iso-3. *Fly (Austin)*. 2012;6(2):80-92.

30. Liu X, Jian X, Boerwinkle E. dbNSFP: a lightweight database of human non-synonymous SNPs and their functional predictions. *Human Mutation*. 2011;32(8):894-899.

31. Liu X, Li C, Mou C, Dong Y, Tu Y. dbNSFP v4: a comprehensive database of transcript-specific functional predictions and annotations for human nonsynonymous and splice-site SNVs. *Genome Med*. 2020;12(1):103.

32. Landrum MJ, Lee JM, Benson M, et al. ClinVar: improving access to variant interpretations and supporting evidence. *Nucleic Acids Res*. 2018;46(D1):D1062-D1067.

33. Stenson PD, Mort M, Ball EV, et al. The Human Gene Mutation Database (HGMD®): optimizing its use in a clinical diagnostic or research setting. *Hum Genet*. 2020;139(10):1197-1207.

34. Jian X, Boerwinkle E, Liu X. In silico prediction of splice-altering single nucleotide variants in the human genome. *Nucleic Acids Res*. 2014;42(22):13534-13544.

35. Xiong HY, Alipanahi B, Lee LJ, et al. RNA splicing. The human splicing code reveals new insights into the genetic determinants of disease. *Science*. 2015;347(6218):1254806.

36. Zook JM, Catoe D, McDaniel J, et al. Extensive sequencing of seven human genomes to characterize benchmark reference materials. *Sci Data*. 2016;3(1):160025.

37. Wei CH, Allot A, Lai PT, et al. PubTator 3.0: an AI-powered literature resource for unlocking biomedical knowledge. *Nucleic Acids Res*. 2024;52(W1):W540-W546.

38. Adzhubei IA, Schmidt S, Peshkin L, et al. A method and server for predicting damaging missense mutations. *Nat Methods*. 2010;7(4):248-249.

39. Milo Rasouly H, Krishna Murthy SB, Vena N, et al. Exome analysis links kidney malformations to developmental disorders and reveals causal genes. *Nat Commun*. 2025;16(1):7290.

40. Neelathi UM, Ullah E, George A, et al. Variants in NR6A1 cause a novel oculo vertebral renal syndrome. *Nat Commun*. 2025;16(1):6111.

41. Merz LM, Kolvenbach CM, Wang C, et al. Trio exome sequencing identifies de novo variants in novel candidate genes in 19.62% of CAKUT families. *Genet Med*. 2025;27(7):101432.

42. Rivera-Munoz EA, Zhao XE, Rosenfeld JA, et al. Clinical exome sequencing efficacy and phenotypic expansions involving non-isolated congenital anomalies of kidney and urinary tract (CAKUT+). *Eur J Hum Genet*. Published online September 6, 2025. doi:10.1038/s41431-025-01929-3

43. Dardas Z, Harrold L, Calame DG, et al. Bi-allelic UGGT1 variants cause a congenital disorder of glycosylation. *Am J Hum Genet*. 2025;112(5):1139-1157.

44. Accogli A, Shakya S, Yang T, et al. Variants in the WDR44 WD40-repeat domain cause a spectrum of ciliopathy by impairing ciliogenesis initiation. *Nat Commun*. 2024;15(1):365.

45. Vaqueiro Graña M, Madariaga L, Gómez-Conde S, Iceta Lizarraga A, Hualde Olascoaga J, Ariceta G. Ultra-rare severe kidney dysplasia mimicking salt-wasting tubulopathy associated with TFCP2L1 gene variants. *Pediatr Nephrol*. Published online June 26, 2025. doi:10.1007/s00467-025-06804-3

46. Amado NG, Nosyreva ED, Thompson D, et al. PIEZO1 loss-of-function compound heterozygous mutations in the rare congenital human disorder Prune Belly Syndrome. *Nat Commun*. 2024;15(1):339.

47. Kesdiren E, Martens H, Brand F, et al. Heterozygous variants in the teashirt zinc finger homeobox 3 (TSHZ3) gene in human congenital anomalies of the kidney and urinary tract. *Eur J Hum Genet*. 2025;33(1):44-55.

48. Mori T, Fujimaru T, Liu C, et al. CFAP47 is implicated in X-linked polycystic kidney disease. *Kidney Int Rep*. 2024;9(12):3580-3591.

49. Elmubarak I, Shril S, Mansour B, et al. Recessive variants in MYO1C as a potential novel cause of proteinuric kidney disease. *Pediatr Nephrol*. 2024;39(10):2939-2945.
